# Predictors of carried ESBL-producing Enterobacterales involvement in ICU-acquired infection: insights from a bicentric retrospective cohort study

**DOI:** 10.64898/2026.07.02.26357103

**Authors:** Caroline Schimpf, Romy Soussan, Paul de Boissieu, Christophe Quesnel, François Philippart

## Abstract

**Rationale:** Infections due to Extended-spectrum β-lactamases-producing *Enterobacterales* (ESBL-PE) require empirical treatment with carbapenems. ESBL-PE carriage is considered as a risk factor for ESBL-PE involvement during ICU infection. Our aim was to determine factors that may predict the actual involvement of ESBL-PE.

**Methods:** A two-periods bicentric ambispective study including ICU ESBL-PE carriers patients from April 2011 to January 2019. All ESBL-PE carriers who developed an infection were analyzed.

**Results:** 6112 patients and 4902 patients were screened during the two periods. 384 and 232 ESBL-PE carriers were identified. Total number of infectious episodes were 146 and 114, respectively. A total of 144 pneumonias, 42 urinary tract infection and 45 digestive infections were studied. An ESBL-PE was involved in 35 (24.3%) episodes of pneumonia, and 44 (37.9%) of extra-pulmonary infections. The most frequent ESBL-PE involved were *K. pneumoniae*, *E. cloacae* and *E. coli*. Similar species and phenotypes were present in colonisation and infection in 29 (82.8%) of pneumonia and in 40 (90.9%) of extra-respiratory infection. Multivariate analysis identified *Klebsiella pneumonia* or *Enterobacter cloacae* carriage as risk factor for ESBL-PE involvement in pneumonia and *E. coli* carriage and detection of ESBL-PE carriage before ICU admission as protective factors.

**Conclusion:** In our study an ESBL-PE involvement is infrequent in pneumonia. A known carriage before ICU admission and *E. coli* carriage are factors associated with the absence of ESBL-PE un the episode of respiratory infection. A confirmation of our findings could lead to a reduction in the empirical use of carbapenems in this population.

## Introduction

Extended-spectrum β-lactamases (ESBL) are enzymes hydrolyzing the amide linkage of the β-lactam ring leading to ineffectiveness of all beta-lactam antibiotics except for cefoxitin and carbepenems. The prevalence of ESBL-producing *Enterobacterales* (ESBL-PE) has gradually increased for nearly three decades (1–5), due both to the emergence of CTX-M type ESBL-PE and to the increase in antibiotic selection pressure favoring the emergence of bacterial strains of reduced susceptibility to antibiotics (1,2,5–7) and became a major health issue in the community (3,5) and in ICU (2,5,8–11). Infections due to ESBL-PE are associated with significant increase in morbidity and mortality (12–15). International guidelines recommend the use of carbapenems for empiric treatment in patients carrying ESBL-PE as known carriage is an independent risk factor of infection involving an ESBL-PE (5,16,17). Moreover, carbapenems appears to be more efficient than piperacillin-tazobactam in treating ESBL-PE infection (18–21), at least for non-urinary tract infections (3). Since carriage is associated with increased involvement of ESBL-PE in subsequent infection (6), especially nosocomial pneumonia (22), and delay in effective antibiotic therapy is associated with a worse prognostic (23–27), carabapenems are favored as first-line treatment (28), particularly in non-urinary tract infections (3) and be maintained for the whole length of treatment in many cases (29). This may lead to an increase in carbapenem exposure in ESBL-PE carriers (28) and to an increased risk of subsequent antibiotic resistance (6).

Conversely the extensive use of carbapenem is associated with an increased risk of selection of multidrug-resistant bacteria, especially carbapenem resistant enterobacterales (30,31). In this context, reducing the unnecessary administration of carbapenem is a central issue. The aim of our study was to measure the involvement of digestive carried ESBL-PE in different ICU-acquired infection.

## Method

### Study settings

We conducted a two-period observational study in two critical care departments (a medical-surgical ICU and a surgical ICU) of two regional university hospitals in Ile-de-France, France. The first period (April 1^st^ 2011 to May 1^st^ 2016) consisted of a retrospective single-center cohort. The second period (February 1^st^ 2017 to January 31^st^ 2019) consisted of a prospective two-centers cohort, extending data collection to a second institution. Both periods were conducted in the same primary center, the second center participated in the prospective period only.

### Patient selection

All patients admitted to our ICU during the defined study period were screened for eligibility. ESBL-PE screening was performed at admission and twice weekly until ICU discharge, according to the same protocol throughout the study period and across both centers. Inclusion criteria were: detection of an ESBL-PE (either on admission or at any subsequent screening point during ICU stay) and occurrence of at least one infectious episode during the ICU stay, with no minimum time interval required between ESBL-PE detection and the infection episode. ESBL-PE carriage could therefore be identified at ICU admission, during ICU stay prior to infection, or concomitantly with the infection episode. Patient with suspected infection that could not be microbiologically documented were excluded.

### Endpoint

The primary endpoint was the proportion ESBL-PE involved in different infections types in ESBL-PE carriers during ICU stay. Secondary endpoints were: (i) identification of factors associated with the absence of ESBL-PE in VAP; (ii) description of ESBL-PE carriage in the study population; (ii) description of all infectious episodes occurring in ESBL-PE carriers during ICU stay.

### Definitions

The same definitions were used during the whole period of the study. Hospital acquired and VAP were defined according to the European (32,33) and American guidelines (34). Microbiological confirmation was obtained by sputum culture (≥10^7^ cfu/ml), tracheal aspirate (≥10^6^ cfu/ml), or, in ventilated patients, bronchoalveolar lavage (≥10^4^ cfu/ml) (34–36).

Urinary tract infection was defined by significant leucocyturia (≥10^4^ /ml) and bacteriuria associated with a clinical and biological inflammation (36). Intraabdominal infections were confirmed by surgical observations and microbiological cultures. Other ICU-acquired infections were diagnosed by the attending physician according to established guidelines (36). Primary bacterial bloodstream infection and central line associated bloodstream infection were defined as in the EUROBACT study (37). Sepsis and septic shock were defined in accordance with the Sepsis-3 definition (38) and acute respiratory distress syndrome by the Berlin definition (38).

Clinical antimicrobial therapy failure was defined as pejorative evolution, with persistence of clinical and/or biological inflammation leading to escalation to broader spectrum antimicrobial therapy, or relapse within 14 days of initiation of initial anti-infective therapy.

### Data collection

For each included patient, the following data were collected : 1) *baseline characteristics*: age, sex, reason for admission, recent abroad travel, chronic organ failure, HIV infection, immunosuppressive treatment, chemotherapy, surgery within the previous 3 months, antimicrobial therapy within the 30 and 90 previous days, Simplified Acute Physiology Score (SAPS II), 2) *infection characteristics*: site, severity, necessity of hemodynamic, respiratory or renal support, association with a bacteremia, empiric and definitive anti-infective therapy, 3)*microbiological data*: preexistent length of known colonization with ESBL-PE, identification and antibiotic microbial susceptibility of ESBL-PE, 4) *outcome*: clinical efficacy of antimicrobial therapy, ICU and hospital length of stay, and survival at ICU discharge, day 28 and hospital discharge.

### Microbiology

All bacterial isolates were sent to the corresponding hospital’s microbiology laboratory. Bacterial identification was performed using the MALDI-TOF mass spectrometry (Andromas system, Paris, France). Antimicrobial susceptibility testing (AST) was performed according to the CA-SFM guidelines (Comité de l’Antibiogramme de la Société Française de Microbiologie) (2011–2014) or EUCAST (2015–2016) by disk diffusion method with Mueller Hinton agar (Biorad, Marnes-la- Coquette, France), using McFarland 0.5 from overnight cultures, followed by incubation at 35°C for 16 to 18 h. Diameters were interpreted accordingly to the breakpoints of CA-SFM or EUCAST guidelines.

All isolates showing reduced susceptibility to ceftazidime or cefotaxime were selected for ESBL detection. ESBL producing bacteria were detected by the double-disk synergy test when bacteria were susceptible to cefoxitin (39). Among AmpC-naturally producing Enterobacteria (*i.e. Enterobacter cloacae, Enterobacter aerogenes, Citrobacter freundii, Morganella morganii*, …) the combination disk method was also applied (cefepime+clavulanate disk *versus* a cefepime disk alone) (40).

### Ethics

The study was carried out in accordance with the Declaration of Helsinki and applicable French law. It was approved by our institutional review board (GERM; IRB n°00012157. Reference 302). Patients were informed about the study and their non-opposition to the collection and use of non-identifying data for research purposes was obtained. Data processing complied with the reference methodology MR-004 of the French Commission Nationale de l’Informatique et des Libertés (French National Data Protection Agency).

### Statistical analysis

Continuous variables were expressed as mean ± standard deviation. Categorical variables were expressed as numbers and percentages.

To identify factors associated with the absence of ESBL-PE involvement in ICU-acquired pneumonia, a logistic regression model was derived from the whole cohort. Candidate variables were selected from clinical relevance, literature consistency and a univariate p-value threshold of less than 0.20. The final model was obtained using a backward stepwise selection procedure, with a retention threshold set at p < 0.05.

Patients with missing data on any variable included in the model were excluded from the analysis. The absence of multicollinearity between retained variables was verified. Model calibration was assessed using the Hosmer-Lemeshow goodness-of-fit test and discrimination using the area under the receiver operating characteristic curve (AUC).

All analyses were performed using SAS software (SAS Institute Inc., Cary, NC, USA).

## Results

### - ESBL-PE colonization

Between April 2011 and January 2019 11 014 patients were admitted in the ICU (6 112 during the first period and 4 902 during the second period). Among them, 618 positive ESBL-PE clinical or screening samples were obtained from 616 unique patients (384 during the first period and 232 during the second period) (**figure 1**).

**Figure 1:**
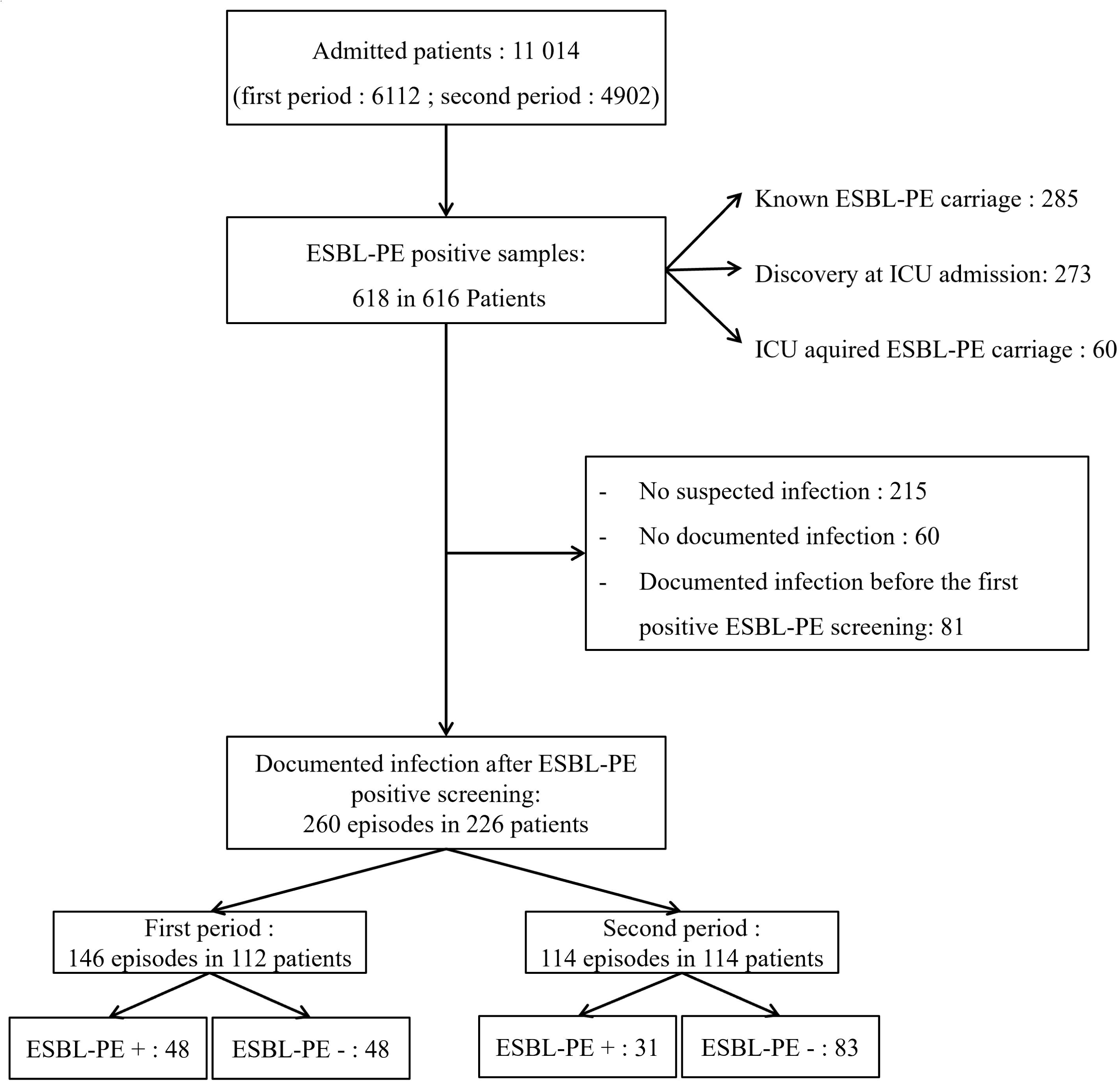
Flow chart All patients included during the two study periods are shown in the flow chart. The two periods are distinguished only in terms of the total number of ICU admissions during each period. ESBL-PE : Extended-spectrum β-lactamases-producing *Enterobacterales;* ICU : Intensive care unit ESBL-PE + : infection involving and ESBL-EP ESBL-PE - : infection not involving and ESBL-EP

ESBL-PE carriage preceded ICU admission in 285 patients (46.3%) ICU acquired ESBL-PE colonization (detected after a negative initial screening) was identified during ICU stay in 60 patients (9.7%) (21 patients (5.4%) in the first period and 39 patients (16.8%) in the second period).

The most frequently isolated ESBL-PE across the entire cohort were *Escherichia coli* (n=398, 64.6%), *Klebsiella pneumoniae* (n=127, 20.6%), and *Enterobacter cloacae* (n=68, 11.0%) (**table 1**).

**Table 1:** ESBL-PE carriage in the total population.

| *Enterobacterale*<br>n (%) | Total<br>(n = 616) |
| --- | --- |
| *Escherichia coli* | 398 (64.6) |
| *Klebsiella pneumoniae* | 127 (20.6) |
| *Enterobacter cloacae* | 68 (11.0) |
| *Klebsiella oxytoca* | 7 (1.1) |
| *Citrobacter freundii* | 2 (0.3) |
| *Citrobacter koserii* | 2 (0.3) |
| *Citrobacter amalonaticus* | 1 (0.2) |
| *Enterobacter aerogenes* | 4 (0.6) |
| *Proteus mirabilis* | 3 (0.5) |
| *Proteus vulgaris* | 3 (0.5) |
| *Serratia marcescens* | 2 (0.3) |

ESBL-EP: Extended-spectrum β-lactamases -producing *Enterobacterales*

### - Infections

Among the 616 ESBL-PE carriers, 226 unique patients (36.7%) developed at least one microbiologically documented infection during their ICU stay, yielding a total of 260 infectious episodes. The primary sites of infections were pneumonia (n=144, 54.9%), digestive infection in (n=45, 17.2%), and urinary tract infection (n=42, 16.0%) (**table 2**). Patients characteristics of ESBL-EP and non-ESBL-PE infected groups is provided in **table 3**. An ESBL-PE was involved in infectious episode in 79 (30.2%) respectively (**table 2, table 3 and supplementary table 1 and 2**).

**Table 2:** Repartition of infections among ESBL-PE carriers.

| Infection location | ESBL-PE +<br>(n = 79)<br>n (%) | ESBL-PE –<br>(n = 181)<br>n (%) |
| --- | --- | --- |
| Pneumonia | 35 (44.3) | 109 (60.2) |
| Urinary tract infection | 24 (30.4) | 18 (9.9) |
| Digestive infection | 15 (19.0) | 30 (12.7) |
| Primary bacteremia / CRI | 1 (1.3) | 12 (3.9) |
| Other | 4 (5.1) | 12 (14.4) |
| First infection episode | 51 (64.6) | 165 (91.2) |
| Second or third infection episode | 28 (35.4) | 18 (9.9) |
| ESBL-PE involved |  |  |
| - *E. coli* | 39 (49.4) | - |
| - *K. pneumoniae* | 22 (27.8) |  |
| - *E. cloacae* | 17 (21.5) |  |
| - *C. freundii* | 1 (1.3) |  |

Other included pleurisy, catheter-related, spondylodiscitis, osteomyelitis, endocarditis, pericarditis, skin infection, surgical site infection.
CRI: Catheter-related infection
ESBL-EP: Extended-spectrum β-lactamases -producing *Enterobacterales*
ESBL-EP+: infection involving and ESBL-EP
ESBL-EP-: infection not involving and ESBL-EP

**Table 3:** infections in the population of ESBL-PE carriers: whole population synthesis.

|  | Total<br>population<br>(n=260) | ESBL-PE<br>infection<br>(n=79) | non ESBL-PE<br>infection<br>(n=181) |
| --- | --- | --- | --- |
| **Demographic data** |  |  |  |
| Age, mean +/- SD | 67.5 (14.7) | 67.0(13.4) | 67.7 (15.1) |
| sex (F), n(%) | 71 (27.1) | 24 (30.4) | 47 (25.7) |
| BMI, mean +/- SD | 25.9 (7.4) | 25.5 ( 7.6) | 26.0 (7.4) |
| Smoker (yes), n (%) | 39 (14.9) | 11 (13.9) | 28 (15.3) |
| Alcohol abuse (yes), n (%) | 22 (8.4) | 5 (6.3) | 17 (9.3) |
| Heart failure, n (%) | 42 (16.0) | 12 (15.2) | 30 (16.4) |
| COPD, n (%) | 41 (15.6) | 5 (6.3) | 36 (19.7) |
| Renal failure, n (%) | 8 (3.1) | 3 ( 3.8) | 5 (2.7) |
| Diabetes mellitus, n (%) | 55 (21.0) | 19 (24.1) | 36 (19.7) |
| Organ transplantation, n (%) | 4 (1.5) | 3 (3.8) | 1 (0.5) |
| Cancer/hematologic malignancy, n (%) | 55 (21.0) | 18 (22.8) | 37 (20.2) |
| AIDS, n (%) | 3 (1.1) | 2 (2.5) | 1 (0.5) |
| Immunosuppressive treatment, n (%) | 32 (12.2) | 8 (10.1) | 24 (13.1) |
| SAPS II, mean +/- SD | 53.8 (19.9) | 56.8 (20.8) | 52.6 (19.4) |
| **Patient origin** |  |  |  |
| Home / Emergency department | 101 (38.5) | 24 (30.4) | 77 (42.1) |
| rehabilitation | 10 (3.8) | 1 (1.3) | 9 (4.9) |
| Ward | 136 (51.9) | 46 (58.2) | 90 (49.2) |
| Foreign country (ward) | 18 (6.9) | 5 (6.3) | 13 (7.1) |
| **Type of admisison** |  |  |  |
| Medical | 167 (63.7) | 51 (64.6) | 116 (63.4) |
| Emergency surgery | 69 (26.3) | 23 (29.1) | 46 (25.1) |
| Planned Surgery | 26 (9.9) | 5 (6.3) | 21 (11.5) |
| **During ICU stay** |  |  |  |
| Vasopressors, n (%) | 116 (44.3) | 37 (46.8) | 79 (43.2) |
| Dialysis, n (%) | 39 (14.9) | 10 (12.6) | 29 (15.8) |
| Invasive mechanical ventilation | 200 (76.3) | 68 (86.1) | 132 (72.1) |
| ARDS | 27 (10.3) | 8 (10.1) | 19 (10.4) |
| prone position | 19 (7.3) | 8 (10.1) | 11 (6.0) |

### - Pneumonias

A total of 144 pneumonia episodes were diagnosed among 128 patients (16 developed 2 episodes) (**table 4**). Ventilator-associated pneumonia (VAP) accounted for 91.4% (32/35) of the ESBL-PE associated episodes and 89.0% (97/109) of the ESBL-PE free infections (table 4). Overall, pneumonia episodes were complicated by acute respiratory distress syndrome (ARDS) in 22 cases (15.3%), septic shock in 78 cases (54.2%) and acute renal failure requiring dialysis in 26 cases (18.1%).

**Table 4:** Characteristics of patients who developed a pneumonia during ICU stay.

|  | Total population |  |
| --- | --- | --- |
|  | ESBL-PE infection (n=35) | non ESBL-PE infection (n=109) |
| **Demographic data** |  |  |
| Age, mean +/- SD | 64.2 (11.8) | 68.0 (15.1) |
| sex (F) , n(%) | 18 (51.4) | 52 (47.7) |
| BMI, mean +/- SD | 24.7 (5.7) | 24.8 (6.2) |
| Current or former smoker (yes), n (%) | 13 (37.1) | 49 (45.0) |
| Alcohol abuse (yes), n (%) | 4 (11.4) | 9 (8.3) |
| Heart failure, n (%) | 3 (8.6) | 14 (12.8) |
| COPD, n (%) | 2 (5.7) | 24 (22.0) |
| Renal failure, n (%) | 1 (2.9) | 2 (1.8) |
| Diabetes mellitus, n (%) | 5 (14.3) | 19 (17.4) |
| Organ transplantation, n (%) | 2 (5.7) | 0 |
| Cancer/hematologic malignancy, n (%) | 6 (17.1) | 17 (15.6) |
| AIDS, n (%) | 1 (2.9) | 2 (1.8) |
| Immunosuppressive treatment, n (%) | 1 (2.9) | 7 (6.4) |
| SAPS II, mean +/- SD | 55.4 (22.6) | 55.1 (21.1) |
| **Previous antibiotic therapy** |  |  |
| Within 30 days | 14 (40.0) | 38 (45.0) |
| Within 90 days | 15 (42.9) | 40 (36.7) |
| **Patient origin** |  |  |
| Home / Emergency department | 14 (40.0) | 45 (41.3) |
| rehabilitation | 0 | 0 |
| Ward | 16 (45.7) | 54 (49.5) |
| Other ICU | 4 (11.4) | 6 (5.5) |
| Foreign country (ward) | 2 (5.7) | 6 (5.5) |
| **Type of admission** |  |  |
| Medical | 30 (85.7) | 83 (76.1) |
| Emergency surgery | 5 (14.3) | 24 (22.0) |
| Planned Surgery | 0 | 2 (1.8) |
| **During ICU stay** |  |  |
| Vasopressors, n (%) | 22 (62.9) | 56 (51.4) |
| Dialysis, n (%) | 7 (20.0) | 19 (17.4) |
| Invasive mechanical ventilation | 32 (91.4) | 97 (89.0) |
| ARDS | 6 (17.1) | 16 (14.7) |
| 28-days mortality | 24 (68.6) | 44 (40.4) |
|  | Total population |  |
|  | ESBL-PE<br>infection<br>(n=35) | Non-ESBL-PE<br>infection<br>(n=109) |
| ESBL-PE carried, n (%) |  |  |
| - *Proteus mirabilis* | 0 | 1 (0.9) |
| - *E. coli* | 12 (34.3) | 74 (67.9) |
| - *Klebsiella pneumoniae* | 6 (17.1) | 21 (19.3) |
| - *Klebsiella oxytoca* | 0 | 3 (2.8) |
| - *Enterobacter cloacae* | 17 (48.6) | 6 (5.5) |
| - *Serratia marcescens* | 0 | 0 |
| - *Proteus vulgaris* | 0 | 1 (0.9) |
| - *Kluyvera spp.* | 0 | 1 (0.9) |
| ESBL-PE involved in pneumonia, n (%) |  |  |
| - *E. coli* | 7 (20.0) | - |
| - *Klebsiella pneumoniae* | 18 (51.4) |  |
| - *Citrobacter koserii* | 1 (2.9) |  |
| - *Enterobacter cloacae* | 8 (22.9) |  |
| - *Citrobacter freundii* | 1 (2.9) |  |
| - Colonization / infection same species, n (%) | 29 (82.8) | - |
| - Colonization / infection different species | 6 (17.2) |  |

An ESBL-PE was involved in the infectious episode in 35 pneumonia episodes (24.3%). The most frequent ESBL-PE pathogens involved were *K. pneumoniae* (n=18, 51.4%), *E. cloacae* (n=8, 22.9%) and *E. coli* (n=7, 20.0%) (**table 4**). In 29 cases (82.8%), the causative ESBL-PE species matched the phenotype and species previously identified in the patient’s screening samples (**table 4**). In the remaining 6 episodes, mismatching species occurred: the carried strain was *E. coli* (n=5) whereas the infectious involved *K. pneumoniae* (n=2), *E. cloacae* (n=2) and *C. freundii* (n=1). The last patient carried a *K. pneumoniae* and developed a pneumonia involving a *C. koserii*. Polymicrobial infections involving an-additional non-ESBL pathogen occured in 10 cases (28.6%) of ESBL-PE associated pneumonias: *Pseudomonas aeruginosa* in 6 (60.0%), *E. coli* (wild type) in 2 (20.0%), *E. cloacae* (wild type) in one (10.0%) and *Streptococcus* spp. in one (10.0%).

Among the 109 ESBL-PE free pneumonias, a total of 131 different pathogens were identified. More than one pathogen was identified in 19 (17.4%) cases. The more frequent pathogen were *Pseudomonas aeruginosa* (n=32, 24.4%), *Escherichia coli* (n=21, 16.0%) and *Staphylococcus aureus* (n=18, 13.7%) (**supplementary tables 3, 4, and 5**).

ESBL-PE associated pneumonia was complicated of septic shock in 22 (62.9%), and acute respiratory distress syndrome in 7 (17.1%) cases. Among ESBL-PE free pneumonia the proportions were 56 (51.4%) and 15 (14.7%) respectively (details in **table 4**).

First episodes of infection occurred 5.9 (+/-7.5) days after ICU admission. The second one occurred 16.1 (+/-18.8) days after ICU admission ESBL-PE was found in 31.8% (34/107) of first respiratory tract infections, in 58.6% (17/29) of second episodes, and 57.1% (4/7) of third episodes.

Total day-28 mortality was 68/128 (53.1%). Considering ESBL-PE associated pneumonia, mortality reached 68.6%.

### - Extra respiratory infections

A total of 116 extra-respiratory infection episodes occurred during the whole study period. Digestive infection (including peritonitis, bile duct infection, and surgical site infection) represent 45 cases (38.8%), urinary tract infection 42 cases (36.2%), primitive bacteremia and catheter-related infection 15 cases (12.9%), cutaneous infection 10 cases (8.6%) (**table 5**). The infection was complicated by septic shock in 61 cases (52.6%) and dialysis requirement in 19 (16.4%).

**Table 5:**
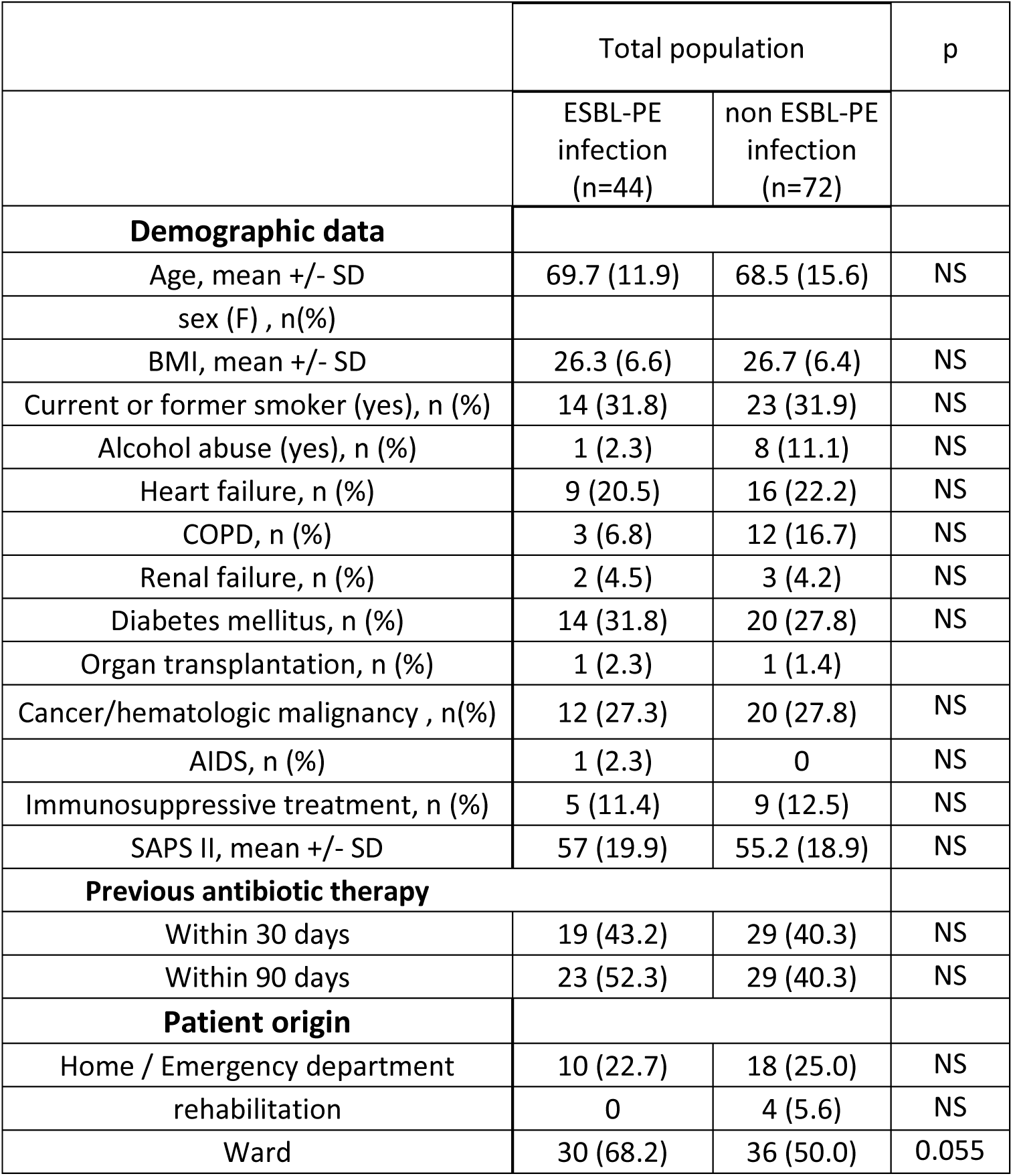
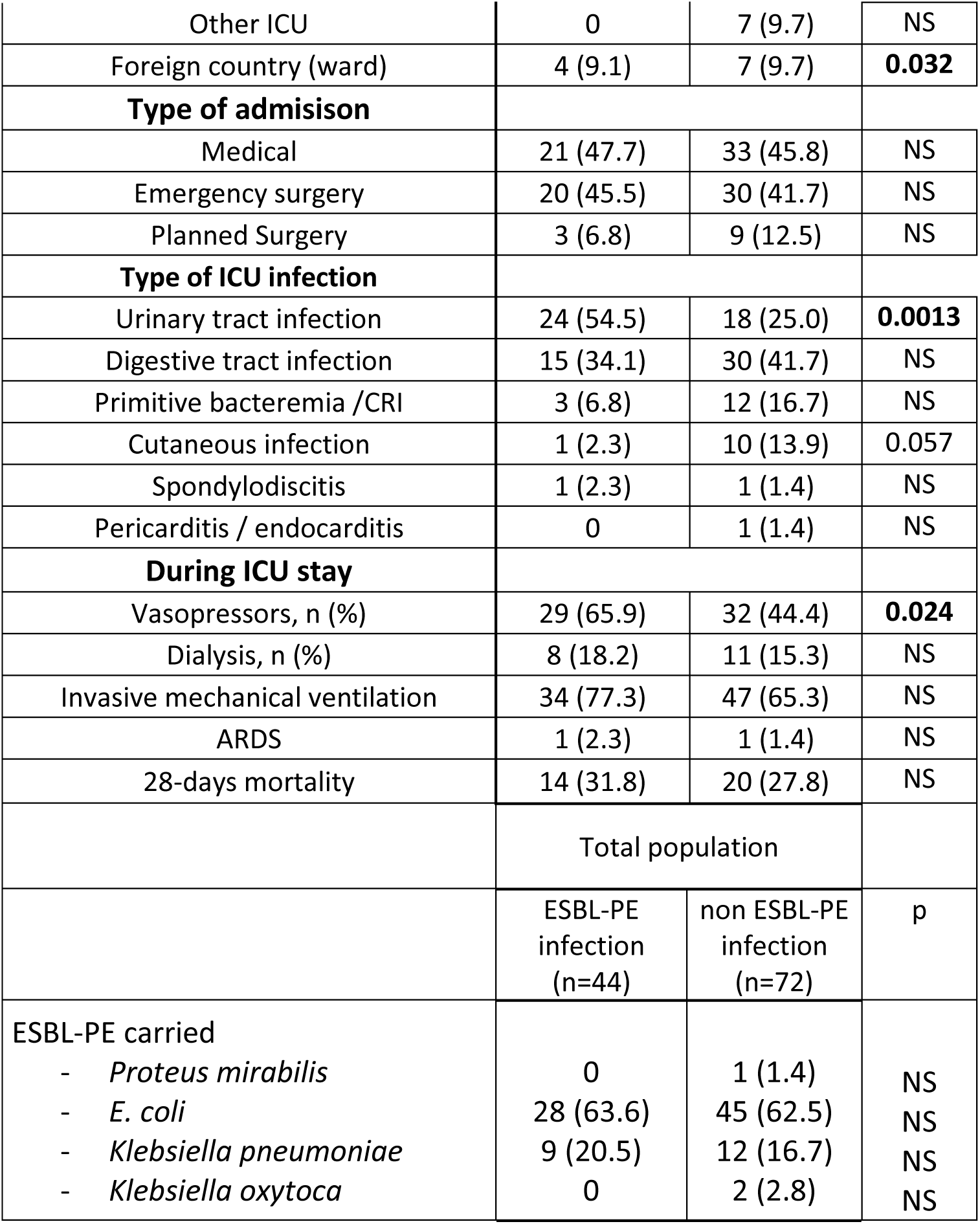
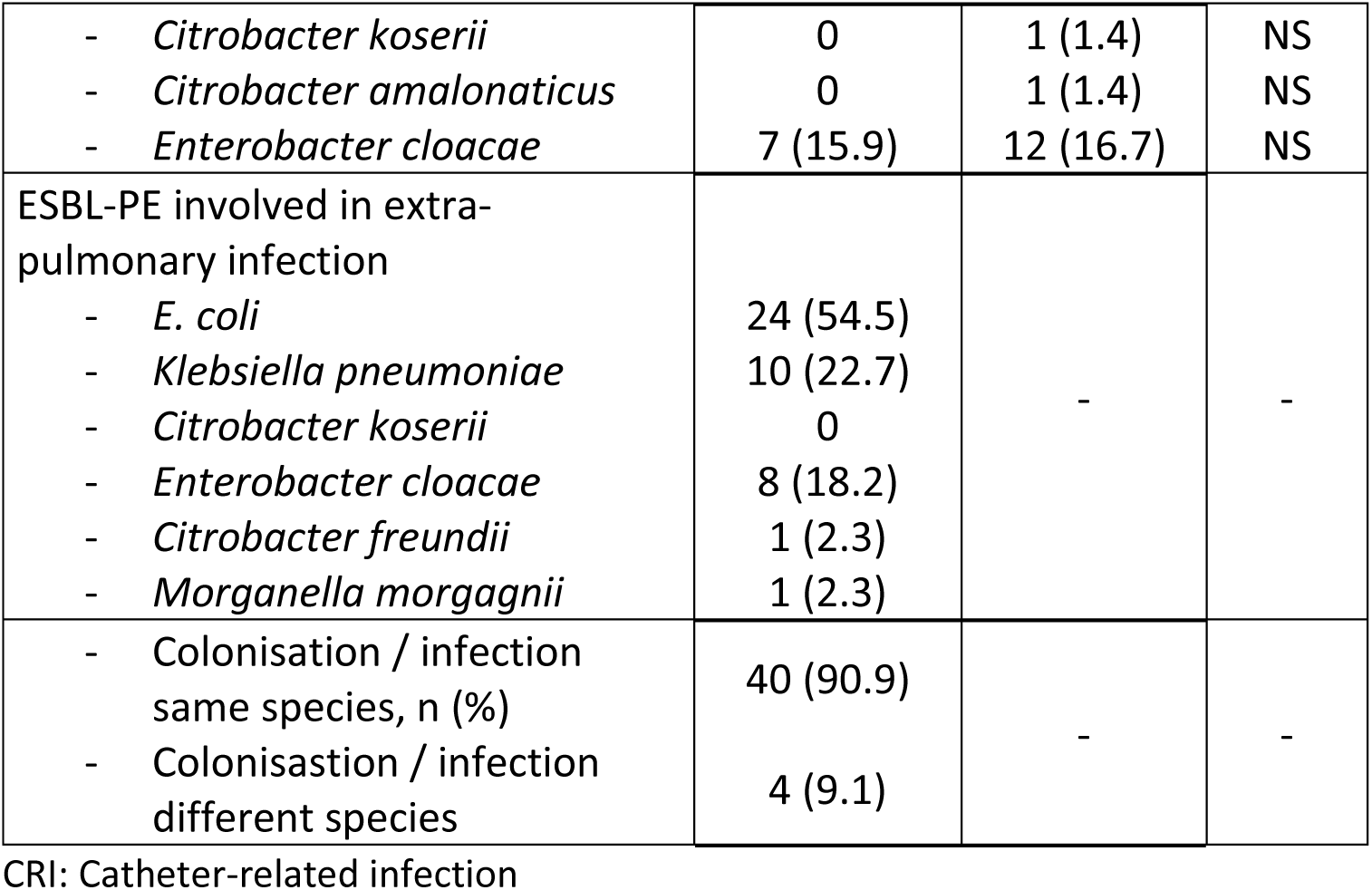
Episodes of extra-pulmonary infection in ESBL-PE carriers.

An ESBL-PE was involved in 44 episodes (37.9%). The most frequent ESBL-PE involved were *E. coli* (n=24, 54.5%), *K. pneumoniae* (n=10, 22.7%) and *E. cloacae* (n=8, 18.2%). A similar species and phenotype to the one found in the portage has been identified in 40 episodes (90.9%). The four remaining situations involved *Citrobacter freundii* (n=1), *Morganella morgagnii* (n=1), *K. pneumoniae* (n=1) and *E. cloacae* (n=1).

ESBLE-PE carriage repartition between the associated and non-ESBL-PE associated infection groups was similar. Repartition of ESBL-PE species between colonisation and infection in the ESBL-PE infected group was also similar.

Among non-ESBL-PE associated infection, most frequent pathogens were *E. coli* (n=17, 18.3%), *P. aeruginosa* (n=13, 14.0%), *S. aureus* (n=9, 9.7%) and *E. faecalis* (n=9, 9.7%) **(supplementary table 6, supplementary table 7)**.

During the whole study period 28-days mortality was 14 (31.8%) in the ESBL-PE infected group and 20 (27.8%) in the non-ESBL-PE associated infection.

### - Risk factor of ESBL-PE involvement during pneumonia in ICU

Considering that Carbapenem is the empiric antibiotic therapy for pneumonia in ESBL-PE carriers, and the need to reduce the use carbapenem due to resistance evolution, we searched potential predictors associated with the absence of ESBL-PE involvement that may prevent the use of such a broad-spectrum antibiotic therapy.

The univariate analysis (**table 6**) revealed that a history of COPD (OR=4.9, 95%CI [1.1-21.8]; p = 0.0376), admission for emergency surgery (OR=3.8, 95%CI [1.1-13.5]; p = 0.1110) or E. coli as carried ESBL-PE (OR=3.5, 95%CI [1.6-7.6]; p = 0.0017) were significantly associated with an increased likehood of the absence or ESBL-PE in the subsequent pneumonia (meaning that these factors could be protective against ESBL-PE pneumonia). Conversely, carrying *K. pneumoniae* (OR=0.3, 95%CI [0.1-0.6]; p = 0.0010) or *E. cloacae* (OR=0.2, 95% CI [0.1-0.6]; p = 0.0029), and identification of carriage during the stay in the ICU (rather than prior to that) (OR=0.2, 95% CI [0.04-0.96]; p = 0.0449) were significantly associated with the odds of an ESBL-free episode.

**Table 6:** Factor associated with the absence of ESLB-PE involvement in pneumonia: Univariate and multivariate analysis.

| Parameters | Univariate Analysis OR [95% CI] | p-value | Multivariate Analysis OR [95% CI] | p-value |
| --- | --- | --- | --- | --- |
| **Demographic data** |  |  |  |  |
| Age (per year) | 1,0 [1,0–1,0] | 0,8 | — | — |
| Sex (Female vs. Male) | 0,9 [0,4–2,1] | 0,8 | — | — |
| BMI (per unit) | 1,0 [0,9–1,1] | 0,9 | — | — |
| Current or former smoker | 1,3 [0,6–2,8] | 0,5 | — | — |
| Alcohol abuse | 0,7 [0,2–2,5] | 0,6 | — | — |
| **Comorbidities** |  |  |  |  |
| Heart failure | 1,2 [0,4–3,9] | 0,8 | — | — |
| COPD | 4,9 [1,1–21,8] | 0,04 | —* | — |
| Diabetes mellitus | 1,1 [0,4–3,1] | 0,8 | — | — |
| Immunocompromission | 1,2 [0,4–3,9] | 0,8 | — | — |
| Cancer / Hematologic malignancy | 1,0 [0,4–2,8] | 1,0 | — | — |
| SAPS II score (at admission) | 1,0 [1,0–1,0] | 0,9 | — | — |
| **Prior antibiotic therapy** |  |  |  |  |
| Within 30 days | 0,7 [0,3–1,5] | 0,4 | — | — |
| Within 90 days | 0,7 [0,3–1,5] | 0,4 | — | — |
| **Patient origin & Admission** |  |  |  |  |
| Ward (vs. Home/Emergency) | 1,3 [0,6–2,7] | 0,8 | — | — |
| Foreign country (ward) | 1,0 [0,2–5,2] | 1,0 | — | — |
| Emergency surgery (vs. Medical) | 3,8 [1,1–13,5] | 0,1 | —* | — |
| Planned Surgery (vs. Medical) | 0,7 [0,1–4,6] | 0,7 | — | — |
| **ESBL-PE Colonization Characteristics** |  |  |  |  |
| Carriage detected during ICU stay (vs. prior to ICU admission) | 0,2 [0,0–1,0] | 0,04 | 0,2 [0,0–0,9] | 0,04 |
| *Escherichia coli* colonization | 3,5 [1,6–7,6] | < 0,01 | — | — |
| *Klebsiella pneumoniae* colonization | 0,3 [0,1–0,6] | < 0,01 | 0,3 [0,1–0,6] | < 0,01 |
| *Enterobacter cloacae* colonization | 0,2 [0,1–0,6] | < 0,01 | 0,1 [0,0–0,3] | < 0,01 |
| **Clinical Presentation** |  |  |  |  |
| HAP (vs. VAP) | 0,9 [0,4–1,9] | 0,8 | — | — |

Values are expressed as Odds Ratios (OR) and 95% Confidence Intervals [95% CI]. Significant p-values are highlighted in bold. —: variable not included in the final multivariable model. \*Variables with an univariable p-value < 0,2 eligible for multivariable step but removed during backward elimination.

In the multivariable logistic regression model (**table 6**), three factors remained independently associated with the absence of ESBL-PE involvement in pneumonia: prior colonization with *E. cloacae* (OR=0.1, 95% CI [0.01-0.3]; p = 0.0004) or *K. pneumoniae* (OR=0.3, 95% CI [0.1-0.6]; p = 0.0029) and identification of ESBL-PE carriage before ICU admission (OR=0.2, 95% CI [0.03-0.9]; p = 0.0421). The model displayed good discriminative capacity, with the area under the receiver operating curve (AUC-ROC) of 0.75 (**figure 2**).

## Discussion

Our results suggest that rectal carriage of ESBL-PE is not systematically associated with an involvement of the resistant pathogen during ICU infections. More specifically whereas their implication is frequent during digestive and urinary tract infection, their role during pneumonia respond to specific criteria, especially during the first episode of infection. The detection of ESBL-PE carriage before ICU admission, and a carriage of ESBL producing *E. coli* suggest an ESBL-PE very unlikely to be involved. Conversely, the presence of *Enterobacter* spp. or *Klebsiella pneumonia* colonization are strongly associated with the involvement of an ESBL-PE in the respiratory tract infection.

To our knowledge very few data are available concerning the potential absence of participation of carried ESBL-PE during ICU-acquired pneumonia. Most studies regarding the implication of ESBL-PE in pneumonia focus on mortality and antibiotic consumption. ESBL-PE incidence is mainly analyzed among bacteremia or urinary and digestive tract infections (5,10,41,42).

In our study, an ESBL-PE is implicated in less than a quarter of the episodes (24.3%) of ICU-acquired pneumonia in the specific population of ESBL-PE carriers. Among ESBL-PE carriers, the proportion of pathogens involved in infections is highly variable function of the case-mix of patients (22,43). Comorbidities, type of admission, initial severity, main admission diagnostic, recent use of antibiotic therapy and delay before infections are all favoring factors (7,22,43,44).

Our findings tend to confirm previous observations (43,45,46). Kevan Razazi *et al.* emphasizing the importance of carried pathogen in the risk of involvement during the first episode of VAP (43,44). In their monocentric retrospective cohort including 111 ICU-acquired pneumonia among ESBL-PE carriers, *Enterobacter* spp. and *Klebsiella pneumoniae* were identified as independent risk factor of ESBL-PE involvement during pneumonia (43). Conversely, the incidence of ESBL producing *E. coli* was ten-fold lower than for *Klebsiella* and *Enterobacter* (1‰ days at risk). On a prospective observation study, Hatem Kallel *et al.* found similar results, as a carried ESBL-PE was involved in 43.2% of ICU-acquired infections (more than 77% of respiratory tract infections), with broad variation function of the pathogen, from 51.2% in *K. pneumoniae* to 5.6% in *E. coli* infections (46). In that study, ESBL-E. coli carriage was associated with a very low positive predictive value (of involvement of an ESBL-E. coli during infection), of 0.056 (46). We found no involvement of *E. coli* in early-onset nosocomial pneumonia, neither in the retrospective part nor in the confirmation cohort in our study. The delay to the first episode of pneumonia was shorter and may have contributed to this result (5.5 *vs* 12 days). However, these results tend to confirm the low implication of *E. coli* in lung infection during the initial days of ICU stay and may lead to a consideration of the possibility of not using carbapenem in probabilistic antibiotic therapy for a first episode occurring before the end of first week of ICU admission.

Many factors may contribute to the low involvement of *E. coli* in early infectious episodes. *Enterobacter* cloacae and *Klebsiella* pneumoniae have specific virulence factor favoring survival, immunity escape, nutrition regulation, especially due to redundant enzymatic system and fine regulation of membrane permeability that improve their fitness and their ability to colonize digestive tract and prevent destruction by host (47–49). These properties may favor secondary infection and proliferation, leading to more severe infections, as clinically observed (43). Similarly, the modification of gut microbiota, associated with specific chronic diseases, ICU stay, therapeutic interventions, mechanical ventilation, may contribute to favor *Klebsiella* and *Enterobacter* proliferation and secondary infection (50,51). The digestive microbiota and its metabolism will contribute to modifying the pulmonary inflammatory response via two main routes: pulmonary recruitment of immune cells from the marrow, whose phenotype has been modified by metabolic mediators from the digestive tract, and direct communication between the digestive and pulmonary microbiota (52). On the same way, the emergence of throat colonization with the same ESBL-PE may be a predictive factor of secondary VAP (45). At last, enrichment of lung microbiome with the low gut bacteria, especially during acute respiratory distress syndrome, play a central role in the risk of secondary nosocomial pneumonia (53). Our results, coherent with previous observation tend to demonstrate that these modifications preferentially involve *Enterobacter* and *Klebsiella* but dynamic analysis of lung and gut microbiota remain to be done.

Beside the modification of the whole microbiome and of the relative abundance of these pathogens, an analysis of their specific abundance may be of interest in the prediction of specific involvement of an ESBL-PE in a subsequent respiratory tract infection (54–56). Moreover, the evaluation of quantitative abundance of ESBL-PE may be of interest both in rectal swab and in higher digestive (54) or respiratory tract (57). This is all the more interesting as the use of prior antibiotic therapy increases the risk of involvement of such bacteria (56). Unfortunately, to date, the analysis of quantitative measurement of ESBL-PE in rectal swab remains unconclusive as a quantitative analysis of seems to predictor of involvement of these bacteria in ICU acquired respiratory tract infection (44) but, on the other hand, a semi-quantitative iterative density measurement of ESBL-PE in rectal swab was associated with ESBL-PE ventilator-associated pneumonia (45). Nonetheless, the correlation between density and later VAP remains insufficient to constitute a major microbiological and resistance predictor in probabilistic antibiotic therapy determination.

As our study was a retrospective analysis of existing data, the biological confirmation of the similarity between the carried ESBL-PE and the strain effectively responsible for the pneumonia remains uncertain. Concordance of species, of antibiograms are suggestive but are still insufficient to confirm with certainty the concordance of the strains. However, a recent paper of Renaud Prevel *et al.* demonstrated in a prospective work that the colonizing strain is indeed the infecting one in their population (7).

Conversely, the regular involvement of the carried ESBL-PE during urinary and digestive tract nosocomial infections is far more expected. The presence of the resistant bacteria in the digestive tract, the risk of perineal and urinary tract colonization and the virulence of *Enterobacterales* are long-established.

Our study has many limitations that may restrict the direct application of its findings. First, the first part of the study was retrospective, introducing potential risks of missing or unrecorded colonization events. To minimize this selection bias, we refrained from calculating the incidence of ESBL-PE carriage among our ICU population, focusing strictly on the risk of infection among documented carriers. As infectious episodes were identified both in ICU files and microbiological laboratory reports, the risk of missing documented infections is expected to be low in the study periods. Second, the regional nature of the study limits the direct external validity of our results to other epidemiological settings. However, the diverse case-mix of the included patients may support the generalizability of these statistical associations to similar tertiary centers. Third, the absence of previous colonization in some patients who developed ESBL-PE pneumonias lead to the question of screening sensibility. Fourth, the observational design precludes any sequential analysis of local microbiota during the ICU stay, meaning that no causal mechanisms regarding microbial dynamics can be inferred from our data. to reinforce the relevance of the observed evolution. Conversely, the observed colonization, especially the predominance of *E. coli*, is fully consistent with established epidemiological data (58). Moreover, by confirming previous observations in a retrospective and a prospective study our work may contribute to the strength of the current observations. This predictive framework represents an exploratory approach toward identifying sub-populations where carbapenem-sparing strategies might be safely evaluated during nosocomial infections.

## Conclusions

In our bicentric, ambispective analysis, the involvement of ESBL-PE in infections varies among ESBL-PE carriers. They are implicated in approximately one third of extrarespiratory cases.

In this cohort, ESBL-PE carriage, alongside *E. cloacae* or *K. pneumoniae* colonization, was independently associated with an increased probability of ESBL-PE involvement in subsequent respiratory infections. Conversely, prior colonization with ESBL-producing E. coli was associated with a critically lower probability of ESBL-PE documented pneumonia.

## Acknowledgement

none

## Authors contribution

Conceptualization: CS,RS,FP; Investigation: CS,RS,CQ, FP; Data curation: CS, RS, PdB; Methodology: PdB; Formal analysis: PdB; Validation : PdB, FP; Supervision: FP; Writing original draft: CS, FP; Writing review and editing : CS, PdB, FP

## Confict of interest

none

## Data availability

Datasets used/analyzed in this study are accessible upon reasonable request to the corresponding author.

---

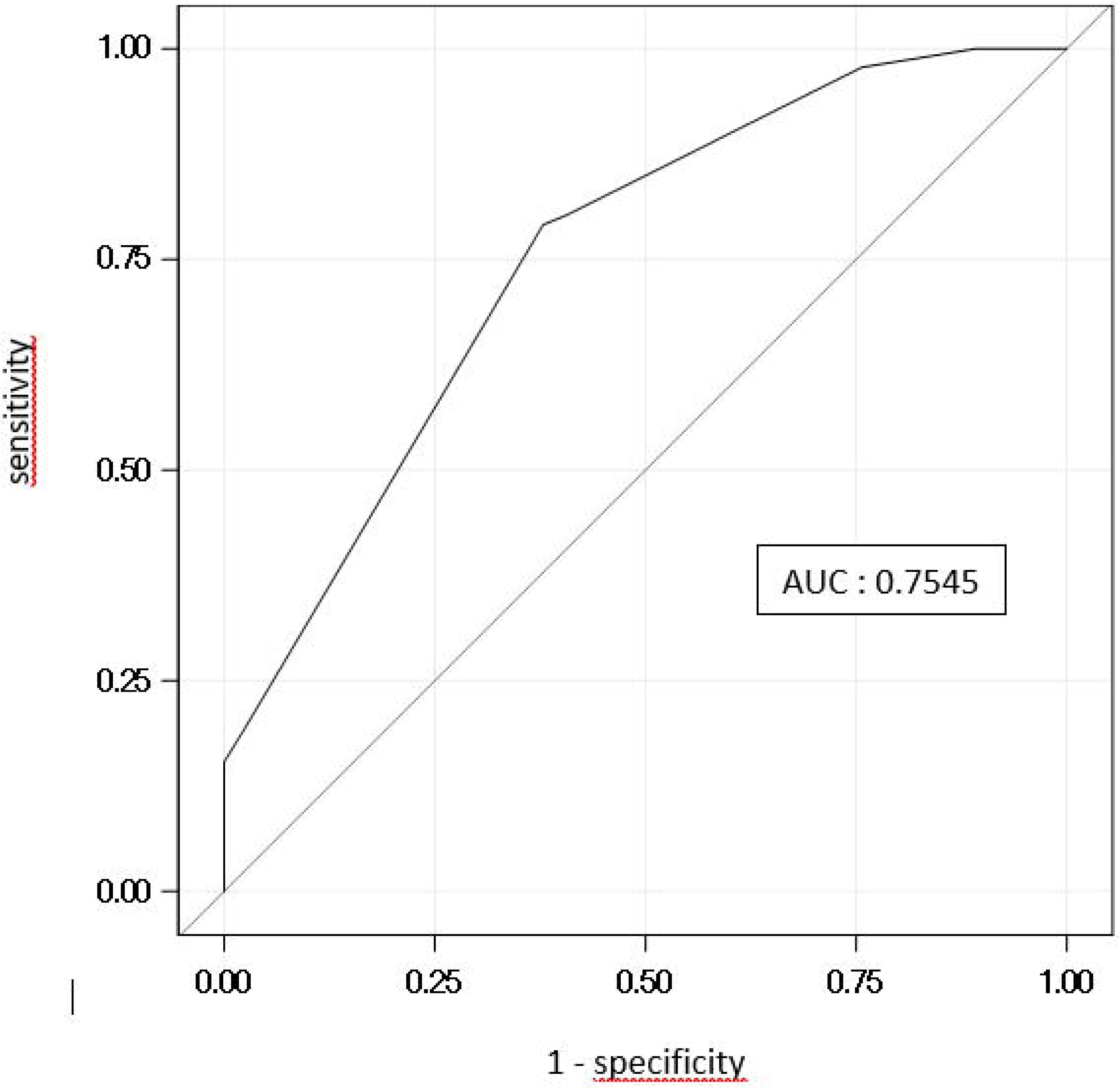

## References

1. Karanika S, Karantanos T, Arvanitis M, Grigoras C, Mylonakis E. Fecal Colonization With Extended-spectrum Beta-lactamase-Producing Enterobacteriaceae and Risk Factors Among Healthy Individuals: A Systematic Review and Metaanalysis. Clin Infect Dis. 1 août 2016;63(3):310-8. doi:10.1093/cid/ciw283 PubMed PMID: 27143671.

2. Sader HS, Farrell DJ, Flamm RK, Jones RN. Antimicrobial susceptibility of Gram-negative organisms isolated from patients hospitalised with pneumonia in US and European hospitals: results from the SENTRY Antimicrobial Surveillance Program, 2009-2012. Int J Antimicrob Agents. avr 2014;43(4):328-34. doi:10.1016/j.ijantimicag.2014.01.007 PubMed PMID: 24630306.

3. Tamma PD, Aitken SL, Bonomo RA, Mathers AJ, van Duin D, Clancy CJ. Infectious Diseases Society of America 2022 Guidance on the Treatment of Extended-Spectrum β-lactamase Producing Enterobacterales (ESBL-E), Carbapenem-Resistant Enterobacterales (CRE), and Pseudomonas aeruginosa with Difficult-to-Treat Resistance (DTR-P. aeruginosa). Clin Infect Dis. 25 août 2022;75(2):187-212. doi:10.1093/cid/ciac268 PubMed PMID: 35439291; PubMed Central PMCID: PMC9890506.

4. Jernigan JA, Hatfield KM, Wolford H, Nelson RE, Olubajo B, Reddy SC, et al. Multidrug-Resistant Bacterial Infections in U.S. Hospitalized Patients, 2012-2017. N Engl J Med. 2 avr 2020;382(14):1309-19. doi:10.1056/NEJMoa1914433 PubMed PMID: 32242356; PubMed Central PMCID: PMC10961699.

5. Adler A, Katz DE, Marchaim D. The Continuing Plague of Extended-spectrum β-lactamase-producing Enterobacteriaceae Infections. Infect Dis Clin North Am. 2016;30(2):347-75. doi:10.1016/j.idc.2016.02.003 PubMed PMID: 27208763.

6. Detsis M, Karanika S, Mylonakis E. ICU Acquisition Rate, Risk Factors, and Clinical Significance of Digestive Tract Colonization With Extended-Spectrum Beta-Lactamase-Producing Enterobacteriaceae: A Systematic Review and Meta-Analysis. Crit Care Med. avr 2017;45(4):705-14. doi:10.1097/CCM.0000000000002253 PubMed PMID: 28157141.

7. Prevel R, Boyer A, M’Zali F, Cockenpot T, Lasheras A, Dubois V, et al. Extended spectrum beta-lactamase producing Enterobacterales faecal carriage in a medical intensive care unit: low rates of cross-transmission and infection. Antimicrob Resist Infect Control. 2019;8:112. doi:10.1186/s13756-019-0572-9 PubMed PMID: 31333839; PubMed Central PMCID: PMC6617905.

8. Pitout JDD, Laupland KB. Extended-spectrum beta-lactamase-producing Enterobacteriaceae: an emerging public-health concern. Lancet Infect Dis. mars 2008;8(3):159-66. doi:10.1016/S1473-3099(08)70041-0 PubMed PMID: 18291338.

9. Chong Y, Yakushiji H, Ito Y, Kamimura T. Clinical and molecular epidemiology of extended-spectrum β-lactamase-producing Escherichia coli and Klebsiella pneumoniae in a long-term study from Japan. Eur J Clin Microbiol Infect Dis. janv 2011;30(1):83-7. doi:10.1007/s10096-010-1057-1 PubMed PMID: 20859753.

10. Razazi K, Derde LPG, Verachten M, Legrand P, Lesprit P, Brun-Buisson C. Clinical impact and risk factors for colonization with extended-spectrum β-lactamase-producing bacteria in the intensive care unit. Intensive Care Med. nov 2012;38(11):1769-78. doi:10.1007/s00134-012-2675-0 PubMed PMID: 22893223.

11. Houard M, Rouzé A, Ledoux G, Six S, Jaillette E, Poissy J, et al. Relationship between digestive tract colonization and subsequent ventilator-associated pneumonia related to ESBL-producing Enterobacteriaceae. PLoS ONE. 2018;13(8):e0201688. doi:10.1371/journal.pone.0201688 PubMed PMID: 30089150; PubMed Central PMCID: PMC6082537.

12. Kola A, Maciejewski O, Sohr D, Ziesing S, Gastmeier P. Clinical impact of infections caused by ESBL-producing E. coli and K. pneumoniae. Scand J Infect Dis. 2007;39(11-12):975-82. doi:10.1080/00365540701466140 PubMed PMID: 17852950.

13. Lee MK, Kim SH, Yong SJ, Shin KC, Park HC, Choi J, et al. Clinical and microbiological features of patients admitted to the intensive care unit with nursing and healthcare-associated pneumonia. J Int Med Res. avr 2015;43(2):236-49. doi:10.1177/0300060514551188 PubMed PMID: 25563575.

14. Palmer HR, Palavecino EL, Johnson JW, Ohl CA, Williamson JC. Clinical and microbiological implications of time-to-positivity of blood cultures in patients with Gram-negative bacilli bacteremia. Eur J Clin Microbiol Infect Dis. juill 2013;32(7):955-9. doi:10.1007/s10096-013-1833-9 PubMed PMID: 23397233.

15. Schwaber MJ, Carmeli Y. Mortality and delay in effective therapy associated with extended-spectrum beta-lactamase production in Enterobacteriaceae bacteraemia: a systematic review and meta-analysis. J Antimicrob Chemother. nov 2007;60(5):913-20. doi:10.1093/jac/dkm318 PubMed PMID: 17848376.

16. Kalil AC, Metersky ML, Klompas M, Muscedere J, Sweeney DA, Palmer LB, et al. Executive Summary: Management of Adults With Hospital-acquired and Ventilator-associated Pneumonia: 2016 Clinical Practice Guidelines by the Infectious Diseases Society of America and the American Thoracic Society. Clin Infect Dis. 1 sept 2016;63(5):575-82. doi:10.1093/cid/ciw504 PubMed PMID: 27521441; PubMed Central PMCID: PMC4981763.

17. Torres A, Ewig S, Lode H, Carlet J, European HAP working group. Defining, treating and preventing hospital acquired pneumonia: European perspective. Intensive Care Med. janv 2009;35(1):9-29. doi:10.1007/s00134-008-1336-9 PubMed PMID: 18989656.

18. Gardner A, Nieberg P, Sakoulas G, Wong-Beringer A. Carbapenem de-escalation as an antimicrobial stewardship strategy: a narrative review. JAC-Antimicrobial Resistance. 1 avr 2025;7(2):dlaf022. doi:10.1093/jacamr/dlaf022

19. Munch MW, Granholm A, Jonsson AB, Sjövall F, Helleberg M, Hertz FB, et al. Piperacillin/tazobactam versus carbapenems in patients with severe bacterial infections: A systematic review with meta-analysis. Acta Anaesthesiol Scand. août 2023;67(7):853-68. doi:10.1111/aas.14239 PubMed PMID: 36919866.

20. Tamma PD, Han JH, Rock C, Harris AD, Lautenbach E, Hsu AJ, et al. Carbapenem therapy is associated with improved survival compared with piperacillin-tazobactam for patients with extended-spectrum β-lactamase bacteremia. Clin Infect Dis. 1 mai 2015;60(9):1319-25. doi:10.1093/cid/civ003 PubMed PMID: 25586681; PubMed Central PMCID: PMC4462658.

21. Hareza DA, Cosgrove SE, Simner PJ, Harris AD, Bergman Y, Conzemius R, et al. Is Carbapenem Therapy Necessary for the Treatment of Non-CTX-M Extended-Spectrum β-Lactamase-Producing Enterobacterales Bloodstream Infections? Clin Infect Dis. 15 mai 2024;78(5):1103-10. doi:10.1093/cid/ciad703 PubMed PMID: 37972276; PubMed Central PMCID: PMC11093655.

22. Carbonne H, Le Dorze M, Bourrel AS, Poupet H, Poyart C, Cambau E, et al. Relation between presence of extended-spectrum β-lactamase-producing Enterobacteriaceae in systematic rectal swabs and respiratory tract specimens in ICU patients. Ann Intensive Care. déc 2017;7(1):13. doi:10.1186/s13613-017-0237-x PubMed PMID: 28155050; PubMed Central PMCID: PMC5289933.

23. Muscedere JG, Shorr AF, Jiang X, Day A, Heyland DK, Canadian Critical Care Trials Group. The adequacy of timely empiric antibiotic therapy for ventilator-associated pneumonia: an important determinant of outcome. J Crit Care. juin 2012;27(3):322.e7-14. doi:10.1016/j.jcrc.2011.09.004 PubMed PMID: 22137378.

24. Martin-Loeches I, Torres A, Rinaudo M, Terraneo S, de Rosa F, Ramirez P, et al. Resistance patterns and outcomes in intensive care unit (ICU)-acquired pneumonia. Validation of European Centre for Disease Prevention and Control (ECDC) and the Centers for Disease Control and Prevention (CDC) classification of multidrug resistant organisms. J Infect. mars 2015;70(3):213-22. doi:10.1016/j.jinf.2014.10.004 PubMed PMID: 25445887.

25. Loh LC, Nor Izran Hanim Bt Abdul Samad null, Rosdara Masayuni Bt Mohd Sani null, Raman S, Thayaparan T, Kumar S. Hospital Outcomes of Adult Respiratory Tract Infections with Extended-Spectrum B-Lactamase (ESBL) Producing Klebsiella Pneumoniae. Malays J Med Sci. juill 2007;14(2):36-40. PubMed PMID: 22993489; PubMed Central PMCID: PMC3442624.

26. Cheng WL, Hsueh PR, Lee CC, Li CW, Li MJ, Chang CM, et al. Bacteremic pneumonia caused by extended-spectrum beta-lactamase-producing Escherichia coli and Klebsiella pneumoniae: Appropriateness of empirical treatment matters. J Microbiol Immunol Infect. avr 2016;49(2):208-15. doi:10.1016/j.jmii.2014.05.003 PubMed PMID: 25070279.

27. Iregui M, Ward S, Sherman G, Fraser VJ, Kollef MH. Clinical importance of delays in the initiation of appropriate antibiotic treatment for ventilator-associated pneumonia. Chest. juill 2002;122(1):262-8. PubMed PMID: 12114368.

28. Barbier F, Pommier C, Essaied W, Garrouste-Orgeas M, Schwebel C, Ruckly S, et al. Colonization and infection with extended-spectrum β-lactamase-producing Enterobacteriaceae in ICU patients: what impact on outcomes and carbapenem exposure? J Antimicrob Chemother. avr 2016;71(4):1088-97. doi:10.1093/jac/dkv423 PubMed PMID: 26755492.

29. Pilmis B, Delory T, Groh M, Weiss E, Emirian A, Lecuyer H, et al. Extended-spectrum beta-lactamase-producing Enterobacteriaceae (ESBL-PE) infections: are carbapenem alternatives achievable in daily practice? Int J Infect Dis. oct 2015;39:62-7. doi:10.1016/j.ijid.2015.08.011 PubMed PMID: 26327124.

30. McLaughlin M, Advincula MR, Malczynski M, Qi C, Bolon M, Scheetz MH. Correlations of antibiotic use and carbapenem resistance in enterobacteriaceae. Antimicrob Agents Chemother. oct 2013;57(10):5131-3. doi:10.1128/AAC.00607-13 PubMed PMID: 23836188; PubMed Central PMCID: PMC3811461.

31. Meyer E, Schwab F, Schroeren-Boersch B, Gastmeier P. Dramatic increase of third-generation cephalosporin-resistant E. coli in German intensive care units: secular trends in antibiotic drug use and bacterial resistance, 2001 to 2008. Crit Care. 2010;14(3):R113. doi:10.1186/cc9062 PubMed PMID: 20546564; PubMed Central PMCID: PMC2911759.

32. Torres A, Niederman MS, Chastre J, Ewig S, Fernandez-Vandellos P, Hanberger H, et al. International ERS/ESICM/ESCMID/ALAT guidelines for the management of hospital-acquired pneumonia and ventilator-associated pneumonia: Guidelines for the management of hospital-acquired pneumonia (HAP)/ventilator-associated pneumonia (VAP) of the European Respiratory Society (ERS), European Society of Intensive Care Medicine (ESICM), European Society of Clinical Microbiology and Infectious Diseases (ESCMID) and Asociación Latinoamericana del Tórax (ALAT). Eur Respir J. sept 2017;50(3). doi:10.1183/13993003.00582-2017 PubMed PMID: 28890434.

33. Leone M, Bouadma L, Bouhemad B, Brissaud O, Dauger S, Gibot S, et al. Brief summary of French guidelines for the prevention, diagnosis and treatment of hospital-acquired pneumonia in ICU. Annals of Intensive Care. 3 nov 2018;8(1):104. doi:10.1186/s13613-018-0444-0

34. Kalil AC, Metersky ML, Klompas M, Muscedere J, Sweeney DA, Palmer LB, et al. Management of Adults With Hospital-acquired and Ventilator-associated Pneumonia: 2016 Clinical Practice Guidelines by the Infectious Diseases Society of America and the American Thoracic Society. Clin Infect Dis. 1 sept 2016;63(5):e61-111. doi:10.1093/cid/ciw353 PubMed PMID: 27418577; PubMed Central PMCID: PMC4981759.

35. American Thoracic Society, Infectious Diseases Society of America. Guidelines for the management of adults with hospital-acquired, ventilator-associated, and healthcare-associated pneumonia. Am J Respir Crit Care Med. 15 févr 2005;171(4):388-416. doi:10.1164/rccm.200405-644ST PubMed PMID: 15699079.

36. Calandra T, Cohen J, International Sepsis Forum Definition of Infection in the ICU Consensus Conference. The international sepsis forum consensus conference on definitions of infection in the intensive care unit. Crit Care Med. juill 2005;33(7):1538-48. PubMed PMID: 16003060.

37. Tabah A, Koulenti D, Laupland K, Misset B, Valles J, Bruzzi de Carvalho F, et al. Characteristics and determinants of outcome of hospital-acquired bloodstream infections in intensive care units: the EUROBACT International Cohort Study. Intensive Care Med. 1 déc 2012;38(12):1930-45. doi:10.1007/s00134-012-2695-9

38. Singer M, Deutschman CS, Seymour CW, Shankar-Hari M, Annane D, Bauer M, et al. The Third International Consensus Definitions for Sepsis and Septic Shock (Sepsis-3). JAMA. 23 févr 2016;315(8):801-10. doi:10.1001/jama.2016.0287 PubMed PMID: 26903338; PubMed Central PMCID: PMC4968574.

39. Jarlier V, Nicolas MH, Fournier G, Philippon A. Extended broad-spectrum beta-lactamases conferring transferable resistance to newer beta-lactam agents in Enterobacteriaceae: hospital prevalence and susceptibility patterns. Rev Infect Dis. août 1988;10(4):867-78. PubMed PMID: 3263690.

40. Mohanty S, Gaind R, Ranjan R, Deb M. Use of the cefepime-clavulanate ESBL Etest for detection of extended-spectrum beta-lactamases in AmpC co-producing bacteria. J Infect Dev Ctries. 21 nov 2009;4(1):24-9. PubMed PMID: 20130375.

41. Nasa P, Juneja D, Singh O, Dang R, Singh A. An observational study on bloodstream extended-spectrum beta-lactamase infection in critical care unit: incidence, risk factors and its impact on outcome. Eur J Intern Med. mars 2012;23(2):192-5. doi:10.1016/j.ejim.2011.06.016 PubMed PMID: 22284253.

42. Vodovar D, Marcadé G, Rousseau H, Raskine L, Vicaut E, Deye N, et al. Predictive factors for extended-spectrum beta-lactamase producing Enterobacteriaceae causing infection among intensive care unit patients with prior colonization. Infection. août 2014;42(4):743-8. doi:10.1007/s15010-014-0619-z PubMed PMID: 24728816.

43. Razazi K, Mekontso Dessap A, Carteaux G, Jansen C, Decousser JW, de Prost N, et al. Frequency, associated factors and outcome of multi-drug-resistant intensive care unit-acquired pneumonia among patients colonized with extended-spectrum β-lactamase-producing Enterobacteriaceae. Ann Intensive Care. déc 2017;7(1):61. doi:10.1186/s13613-017-0283-4 PubMed PMID: 28608133; PubMed Central PMCID: PMC5468364.

44. Bay P, Woerther PL, Fihman V, Gendreau S, Labedade P, Gaillet A, et al. Relative faecal abundance to predict extended-spectrum β-lactamase-producing Enterobacterales related ventilator-associated pneumonia. Ann Intensive Care. 20 mars 2025;15(1):34. doi:10.1186/s13613-025-01456-w PubMed PMID: 40113731; PubMed Central PMCID: PMC11925845.

45. Andremont O, Armand-Lefevre L, Dupuis C, de Montmollin E, Ruckly S, Lucet JC, et al. Semi-quantitative cultures of throat and rectal swabs are efficient tests to predict ESBL-Enterobacterales ventilator-associated pneumonia in mechanically ventilated ESBL carriers. Intensive Care Med. juin 2020;46(6):1232-42. doi:10.1007/s00134-020-06029-y PubMed PMID: 32313993; PubMed Central PMCID: PMC7222166.

46. Kallel H, Houcke S, Resiere D, Court T, Roncin C, Raad M, et al. Prior Carriage Predicts Intensive Care Unit Infections Caused by Extended-Spectrum Beta-Lactamase-Producing Enterobacteriaceae. Am J Trop Med Hyg. 10 janv 2022;106(2):525-31. doi:10.4269/ajtmh.20-1436 PubMed PMID: 35008044; PubMed Central PMCID: PMC8832904.

47. Davin-Regli A, Pagès JM. Enterobacter aerogenes and Enterobacter cloacae; versatile bacterial pathogens confronting antibiotic treatment. Front Microbiol. 2015;6:392. doi:10.3389/fmicb.2015.00392 PubMed PMID: 26042091; PubMed Central PMCID: PMC4435039.

48. Bachman MA, Breen P, Deornellas V, Mu Q, Zhao L, Wu W, et al. Genome-Wide Identification of Klebsiella pneumoniae Fitness Genes during Lung Infection. Gilmore MS, éditeur. mBio. juill 2015;6(3). doi:10.1128/mBio.00775-15

49. Martin RM, Bachman MA. Colonization, Infection, and the Accessory Genome of Klebsiella pneumoniae. Front Cell Infect Microbiol. 2018;8:4. doi:10.3389/fcimb.2018.00004 PubMed PMID: 29404282; PubMed Central PMCID: PMC5786545.

50. Bai Y, Hu Y, Chen X, Hu L, Wu K, Liang S, et al. Comparative metagenome-associated analysis of gut microbiota and antibiotic resistance genes in acute gastrointestinal injury patients with the risk of in-hospital mortality. mSystems. 18 mars 2025;10(3):e0144424. doi:10.1128/msystems.01444-24 PubMed PMID: 40013797; PubMed Central PMCID: PMC11915821.

51. Xu J, Kong X, Li J, Mao H, Zhu Y, Zhu X, et al. Pediatric intensive care unit treatment alters the diversity and composition of the gut microbiota and antimicrobial resistance gene expression in critically ill children. Front Microbiol. 2023;14:1237993. doi:10.3389/fmicb.2023.1237993 PubMed PMID: 38029168; PubMed Central PMCID: PMC10679412.

52. Lloyd CM, Marsland BJ. Lung Homeostasis: Influence of Age, Microbes, and the Immune System. Immunity. 18 avr 2017;46(4):549-61. doi:10.1016/j.immuni.2017.04.005 PubMed PMID: 28423336.

53. Dickson RP, Singer BH, Newstead MW, Falkowski NR, Erb-Downward JR, Standiford TJ, et al. Enrichment of the lung microbiome with gut bacteria in sepsis and the acute respiratory distress syndrome. Nat Microbiol. 18 juill 2016;1(10):16113. doi:10.1038/nmicrobiol.2016.113 PubMed PMID: 27670109; PubMed Central PMCID: PMC5076472.

54. Gorrie CL, Mirceta M, Wick RR, Edwards DJ, Thomson NR, Strugnell RA, et al. Gastrointestinal Carriage Is a Major Reservoir of Klebsiella pneumoniae Infection in Intensive Care Patients. Clin Infect Dis. 15 juill 2017;65(2):208-15. doi:10.1093/cid/cix270 PubMed PMID: 28369261; PubMed Central PMCID: PMC5850561.

55. Pilmis B, Mizrahi A, Péan de Ponfilly G, Philippart F, Bruel C, Zahar JR, et al. Relative faecal abundance of extended-spectrum β-lactamase-producing Enterobacterales and its impact on infections among intensive care unit patients: a pilot study. J Hosp Infect. juin 2021;112:92-5. doi:10.1016/j.jhin.2021.03.022 PubMed PMID: 33794294.

56. Mu S, Xiang H, Wang Y, Wei W, Long X, Han Y, et al. The pathogens of secondary infection in septic patients share a similar genotype to those that predominate in the gut. Crit Care. 24 mars 2022;26(1):68. doi:10.1186/s13054-022-03943-z PubMed PMID: 35331299; PubMed Central PMCID: PMC8944137.

57. Fromentin M, Ricard JD, Roux D. Respiratory microbiome in mechanically ventilated patients: a narrative review. Intensive Care Med. mars 2021;47(3):292-306. doi:10.1007/s00134-020-06338-2 PubMed PMID: 33559707; PubMed Central PMCID: PMC7871139.

58. Pilmis B, Cattoir V, Lecointe D, Limelette A, Grall I, Mizrahi A, et al. Carriage of ESBL-producing Enterobacteriaceae in French hospitals: the PORTABLSE study. J Hosp Infect. mars 2018;98(3):247-52. doi:10.1016/j.jhin.2017.11.022 PubMed PMID: 29222035.

