## Supplementary material for "Predictors of carried ESBL-producing Enterobacterales involvement in ICU-acquired infection: insights from a bicentric retrospective cohort study"

**Antimicrobial therapy during pneumonia in ESBL-PE colonized patients**

Among 78 pneumonias, 69 (88%) received empirical therapy. Empirical therapy included carbapenem in 26 cases (38%), the association of piperacillin and tazobactam in 24 cases (35%), ceftriaxone in 9 (13%), amoxicillin and clavulanate in 7 (10%), piperacillin or cefepime or ceftazidime in one case each. An aminoglycoside was associated in 26 cases (38%) (**Table 5**). Considering ESBL-PE-associated pneumonia, piperacillin and tazobactam was the first line therapy in 7 cases (38%), with an *in vitro* efficiency in 6 cases; imipenem was administered in 9 cases (50%), with no *in vitro* resistance, and an aminoglycoside was associated in 8 cases (42%). In vitro efficiency of amikacin was retained in all cases. Definitive antimicrobial therapy consisted of carbapenems in 11 cases (58%), piperacillin and tazobactam in 6 cases (32%).

Mean length of empirical therapy was 2 days; mean length of antimicrobial therapy was 9.3 days. Clinical failure was observed in one patient despite an *in vitro* efficiency of the administered therapy.

Notably, three of ESBL-PE$^-$ pneumonia patients secondary developed an ESBL-PE$^+$ pneumonia before dying, despite an empiric therapy which always included a carbapenem.

Supplementary table 1: ESBL-PE involved in infection during ICU stay

| *Enterobacterale involved* | First period cohort (n= 384) Infected patients (n=112) Number of episodes (n=146) | | Second period cohort (n= 232) Infected patients (n=114) Number of episodes (n=114) | |
|---|---|---|---|---|
| | Colonization (n= 384) | Infection ESBL-PE+ (48) | Colonization (n= 232) | Infection ESBL-PE+ (31) |
| *Escherichia coli* | 261 (67.9%) | 29 (60.4) | 137 (59%) | 10 (32.3) |
| *Klebsiella pneumoniae* | 73 (19%) | 8 (16.7) | 54 (23.3%) | 14 (45.2) |
| *Enterobacter cloacae* | 38 (9.9%) | 11 (22.9) | 30 (12.9%) | 6 (19.3) |
| *Klebsiella oxytoca* | 4 (1.0%) | - | 3 (1.3%) | - |
| *Citrobacter freundii* | - | - | 2 (0.8%) | 1 (3.2) |
| *Enterobacter aerogenes* | 4 (1.0%) | - | - | - |
| *Proteus mirabilis* | 2 (0.5%) | - | 1 (0.4%) | - |
| *Serratia marcescens* | 2 (0.5%) | - | - | - |
| Others | 0 | - | 5 (16.1) | - |

Supplementary table 2: ESBL-PE carried by patients with non-ESBL-PE-involved infection

| *Enterobacterale involved* | First period cohort (n= 384) Infected patients (n=112) Number of episodes (n=114) | Second period cohort (n= 232) Infected patients (n=114) Number of episodes (n=114) |
|---|---|---|
| | ESBL- (98)* | ESBL- (83) |
| *Escherichia coli* | 71 (71) | 45 (54.2) |
| *Klebsiella pneumoniae* | 15 (15) | 19 (22.9) |
| *Enterobacter cloacae* | 9 (9) | 10 (12.0) |
| *Klebsiella oxytoca* | 1 (1) | 2 (2.4) |
| *Citrobacter freundii* | - | 1 (1.2) |
| *Enterobacter aerogenes* | 3 (3) | - |
| *Proteus mirabilis* | 1 (1) | 1 (1.2) |
| *Serratia marcescens* | 1 (1) | - |

*NB: Two ESBL-PE were carried by two patients: *E. coli* and *K. pneumoniae* in both.

Supplementary table 3: Pathogens associated with non-ESBL-PE pneumonia

| Pathogen | n (%) | | |
|---|---|---|---|
| | Total population | Initial cohort | Second period cohort |
| *Pseudomonas aeruginosa* | 32 (24.4) | 14 (19.4) | 18 (30.4) |
| *Escherichia coli* | 21 (16.0)) | 15 (20.8) | 6 (10.2) |
| *Staphylococcus aureus* | 18 (13.7) | 12 (16.7) | 6 (10.2) |
| Oropharyngeal flora | 10 (7.6) | 4 (5.6) | 6 (10.2) |
| *Klebsiella pneumoniae* | 8 (7.6) | 2 (2.8) | 6 (10.2) |
| *Haemophilus influenzae* | 7 (5.3) | 5 (6.9) | 2 (3.4) |
| *Streptococcus pneumoniae* | 6 (4.6) | 4 (5.6) | 2 (3.4) |
| *Serratia marcescens* | 5 (3.8) | 4 (5.6) | 1 (1.7) |
| *Pneumocystis carinii* | 4 (3.1) | - | 4 (6.8) |
| *Enterococcus faecalis* | 3 (2.3) | (1 (1.4) | 2 (3.4) |
| *Stenotrophomonas Maltophilia* | 3 (2.3) | 1 (1.4) | 2 (3.4) |
| *Proteus mirabilis* | 2 (1.5) | - | 2 (3.4) |
| *Morganella morgagnii* | 2 (1.5) | 1 (1.4) | 1 (1.7) |
| *Streptococcus spp.* | 2 (1.5) | 2 (2.8) | - |
| *Alcaligenes xylosoxidans* | 2 (1.5) | 2 (2.8) | - |
| *Klebsiella oxytoca* | 1 (0.8) | 1 (1.4) | - |
| *Citrobacter koserii* | 1 (0.8) | 1 (1.4) | - |
| *Citrobacter freundii* | 1 (0.8) | 1 (1.4) | - |
| *Hafnia alvei* | 1 (0.8) | 1 (1.4) | - |
| *Acinetobacter baumanii* | 1 (0.8) | - | 1 (1.7) |
| *Pasteurella multocida* | 1 (0.8) | 1 (1.4) | - |

Supplementary table 4: co-pathogen in non ESBL-PE associated pneumonia

| Total population : n (%) | |
|---|---|
| *Pseudomonas aeruginosa* :12 (37.5) | - *Escherichia coli* : 5 (41.7)<br>- *Staphylococcus aureus* : 3 (25.0)<br>- *Enterococcus faecalis* : 1 (8.3)<br>- *Morganella morgagnii* : 1 (8.3)<br>- *Serratia marcescens* : 1 (8.3) |
| *Staphylococcus aureus* : 10 (50) | - *Escherichia coli* : 4 (40.0)<br>- *Pseudomonas aeruginosa* : 3 (30.0)<br>- *Klebsiella pneumoniae* : 1 (10.0)<br>- *Klebsiella oxytoca* : 1 (10.0)<br>- *Morganella morgagnii* : 1 (10.0) |
| *Escherichia coli* : 13 (61.9) | - *Pseudomonas aeruginosa* :5 (38.5)<br>- *Staphylococcus aureus* :4 (30.8)<br>- *Hemophilus influenzae* : 1 (7.7)<br>- *Citrobacter koserii* : 1 (7.7)<br>- *Serratia marcescens* : 1 (7.7)<br>- *Stenotrophomonas maltophilia* : 1 (7.7) |
| *Hemophilus influenzae* : 5 (71.4) | - *Streptococcus pneumoniae* : 4 (80.0)<br>- *Escherichia coli* : 1 (20.0) |
| *Enterococcus faecalis* : 2 (100) | - *Pseudomonas aeruginosa* : 1 (50.0)<br>- *Citrobacter freundii* : 1 (50.0) |
| *Klebsiella pneumoniae* : 3 (37.5) | - *Proteus mirabilis* : 1 (33.3)<br>- *Serratia marcescens* : 1 (33.3)<br>- *Acinetobacter baumanii* : 1 (33.3) |
| *Serratia marcescens* : 4 (50.0) | - *Pseudomonas aeruginosa* : 2 (50.0)<br>- *Escherichia coli* : 1 (25.0)<br>- *Klebsiella pneumoniae* : 1 (25.0) |
| *Streptococcus pneumoniae* : 5 (83.3) | - *Hemophilus influenzae* : 4 (80.0)<br>- *Branhamella cattharalis* : 1 (20.0) |
| *Morganella morgagnii* :2 (66.7) | - *Staphylococcus aureus* : 1 (50.0)<br>- *Pseudomonas aeruginosa* : 1(50.0) |
| *Acinetobacter baumanii* : 1 (100) | - *Klebsiella pneumoniae* : 1 (100) |
| *Stenotrophomonas maltophilia* : 1 (100) | - *Escherichia coli* : 1 (100) |
| *Proteus mirabilis* : 1 (50.0) | - *Klebsiella pneumoniae* : 1 (100) |
| *Citrobacer koserii* : 1 (100) | - *Escherichia coli* : 1 (100) |
| *Branhamella cattharalis* : 1 (50.0) | - *Streptococcus pneumoniae* : 1 (100) |

Supplementary table 5: non-ESBL-PE pathogens involved in non ESBL-PE associated pneumonia, function of ESBL-PE carriage

| Total population | | First period cohort | Second period cohort |
|---|---|---|---|
| Carried ESBL-PE | Pathogen associated pneumonia ; n (%) | Pathogen associated pneumonia ; n (%) | Pathogen associated pneumonia ; n (%) |
| *Escherichia coli* | *Pseudomonas aeruginosa* :21 (21.9)<br>*E. coli* : 17 (17.7)<br>*Staphylococcus aureus* : 15 (15.6)<br>*Haemophilus influenzae* : 7 (7.3)<br>Oropharyngeal flora : 5 (5.2)<br>*Streptococcus pneumoniae* : 5 (5.2)<br>*Klebsiella pneumoniae* : 5 (5.2)<br>*Serratia marcescens* : 4 (4.2)<br>*Enterococcus faecalis* : 3 (3.1)<br>*Streptococcus* spp : 2 (2.1)<br>*Morganella morgagnii* : 2 (2.1)<br>*Alcaligeness xylosoxidans* : 2 (2.1)<br>*Pneumocystis jirovecii* : 2 (2.1)<br>*Proteus mirabilis* : 1 (1.0)<br>*Klebsiella oxytoca*: 1 (1.0)<br>*Citrobacter freundii*: 1 (1.0)<br>*Acinetobacter baumanii* : 1 (1.0)<br>*Stenotrophomonas maltophilia* : 1 (1.0)<br>Pasteurella multocida: 1 (1.0) | *E. coli* :13 (21.3)<br>*Pseudomonas aeruginosa* : 12 (19.7)<br>*Staphylococcus aureus* : 11 (18.0)<br>*Haemophilus influenzae* : 5 (8.2)<br>*Streptococcus pneumoniae* :4 (6.6)<br>*Serratia marcescens* : 3 (4.9)<br>*Streptococcus spp.* : 2 (3.3)<br>*Klebsiella pneumoniae* : 2 (3.3)<br>*Alcaligeness xylosoxidans* : 2 (3.3)<br>Oropharyngeal flora : 1 (1.6)<br>*Enterococcus faecalis* : 1 (1.6)<br>*Klebsiella oxytoca*: 1 (1.6)<br>*Morganella morgagnii* : 1 (1.6)<br>*Citrobacter freundii*: 1 (1.6)<br>Pasteurella multocida: 1 (1.6)<br>*Stenotrophomonas maltophilia* : 1 (1.6) | *Pseudomonas aeruginosa* :9 (25.7)<br>*E. coli* : 4 (11.4)<br>*Staphylococcus aureus* : 4 (11.4)<br>Oropharyngeal flora :4 (11.4)<br>*Klebsiella pneumoniae* : 3 (8/6)<br>*Haemophilus influenzae* : 2 (5.7)<br>*Enterococcus faecalis* : 2 (5.7)<br>*Pneumocystis jirovecii* : 2 (5.7)<br>*Streptococcus pneumoniae* : 1 (2.9)<br>*Proteus mirabilis* : 1 (2.9)<br>*Morganella morgagnii* : 1 (2.9)<br>*Serratia marcescens* : 1 (2.9)<br>*Acinetobacter baumanii* : 1 (2.9) |
| *Klebsiella pneumoniae* | Oropharyngeal flora : 2 (22.2)<br>*E. coli* : 2 (22.2)<br>*Pseudomonas aeruginosa* : 2 (22.2)<br>*Staphylococcus aureus* : 1 (11.1)<br>*Citrobacter koserii* : 1 (11.1)<br>*Serratia marcescens*: 1 (11.1) | *Pseudomonas aeruginosa* : 6 (37.5)<br>*Staphylococcus aureus* : 2 (12.5)<br>*E. coli* :2 (12.5)<br>*Klebsiella pneumoniae* : 2 (12.5)<br>Oropharyngeal flora : 1 (6.25)<br>*Proteus mirabilis* :1 (6.25)<br>*Stenotrophomonas maltophilia* : 1(6.25) | *Pseudomonas aeruginosa* : 8 (32.0)<br>*E. coli* :4 (16.0)<br>Oropharyngeal flora : 3 (12.0)<br>*Staphylococcus aureus* :3 (12.0)<br>*Klebsiella pneumoniae* : 2 (8.0)<br>*Proteus mirabilis* :1 (4.0)<br>*Citrobacter koserii* : 1 (4.0) |

| | | | |
|---|---|---|---|
| | | *Pneumocystis jirovecii* : 1 (6.25) | *Serratia marcescens*: 1 (4.0)<br>*Stenotrophomonas maltophilia* : 1 (4.0)<br>*Pneumocystis jirovecii* : 1 (4.0) |
| *Enterobacter cloacae* | Oropharyngeal flora : 1 (50.0)<br>*Hafnia alvei* : 1 (50.0) | *Pseudomonas aeruginosa* : 3 (60.0)<br>*Klebsiella pneumoniae* : 1 (20.0)<br>*Stenotrophomonas maltophilia* : 1 (20.0) | *Pseudomonas aeruginosa* : 3 (42.9)<br>Oropharyngeal flora : 1 (14.3)<br>*Klebsiella pneumoniae* : 1 (14.3)<br>*Hafnia alvei* : 1 (14.3)<br>*Stenotrophomonas maltophilia* : 1 (14.3) |
| *Klebsiella oxytoca* | - | *Pneumocystis jirovecii : 1* | *Pneumocystis jirovecii : 1* |
| *Proteus vulgaris* | - | *Oropharyngeal flora : 1* | *Oropharyngeal flora : 1* |
| *Kluyvera spp.* | - | *S. pneumoniae : 1* | *S. pneumoniae : 1* |

Supplementary table 6: Pathogens associated with non-ESBL-PE extra-respiratory infections

| Pathogen | n (%) | | |
|---|---|---|---|
| | Total population | First period cohort | Second period cohort |
| *E. coli* | 17 (18.3) | 11 (23.4) | 6 (13.3) |
| *P. aeruginosa* | 13 (14.0) | 6 (12.8) | 7 (15.6) |
| *S. aureus* | 9 (9.7) | 3 (6.4) | 6 (13.3) |
| *E. faecalis* | 9 (9.7) | 5 (10.6) | 4 (8.9) |
| *E. cloacae* | 9 (9.7) | 4 (8.5) | 5 (11.1) |
| *Streptococcus spp.* | 7 (7.5) | 4 (8.5) | 3 (6.7) |
| *Non aureus Staphylococcus* | 5 (5.4) | 2 (4.3) | 3 (6.7) |
| *K. pneumoniae* | 4 (4.3) | 2 (4.3) | 2 (4.4) |
| *S. marcescens* | 4 (4.3) | 1 (2.1) | 3 (6.7) |
| *E. avium* | 2 (2.2) | 1 (2.1) | 1 (2.2) |
| *P. mirabilis* | 2 (2.2) | 2 (4.3) | - |
| *M. morgagnii* | 2 (2.2) | 1 (2.1) | 1 (2.2) |
| *S. pneumoniae* | 1 (1.1) | 1 (2.1) | - |
| *E. faecium* | 1 (1.1) | 1 (2.1) | - |
| *K.oxytoca* | 1 (1.1) | - | 1 (2.2) |
| *E. aerogenes* | 1 (1.1) | 1 (2.1) | - |
| *C. freundii* | 1 (1.1) | - | 1 (2.2) |
| *H. alvei* | 1 (1.1) | - | 1 (2.2) |
| *B. fragilis* | 2 (2.2) | 1 (2.1) | 1 (2.2) |
| *Acinetobacter Iwoffii* | 1 (1.1) | - | 1 (2.2) |
| *Lactobacillus spp.* | 1 (1.1) | 1 (2.1) | - |

Supplementary table 7: non-ESBL-PE pathogens involved in non ESBL-PE associated infection (except pneumonia), function of ESBL-PE carriage

| Total population | | First period cohort | Second period cohort |
|---|---|---|---|
| Carried ESBL-PE | Pathogen associated pneumonia ; n (%) | Pathogen associated pneumonia ; n (%) | Pathogen associated pneumonia ; n (%) |
| *E. coli* | *E. coli* : 14 (25.9)<br>*S. aureus* : 8 (14.8)<br>*P. aeruginosa* : 6 (11.1)<br>*Streptococcus spp.* : 4 (7.4)<br>*E. faecalis* : 4 (7.4)<br>*K. pneumoniae* : 3 (5.6)<br>*E. cloacae* : 3 (5.6)<br>Non aureus *Staphylococcus* : 2 (3.7)<br>*Enterococcus avium* : 2 (3.7)<br>*S. pneumoniae* : 1 (1.9)<br>*P. mirabilis* : 1 (1.9)<br>*Enterobacter aerogenes* : 1 (1.9)<br>*S. marcescens* : 1 (1.9)<br>*C. freundii* : 1 (1.9)<br>*M. morgagnii* : 1 (1.9)<br>*Bacillus fragilis* : 1 (1.9)<br>*Lactobacillus* spp : 1 (1.9) | *E. coli* : 9 (29.0)<br>*P. aeruginosa* : 4 (12.9)<br>*Streptococcus spp.* : 4 (12.9)<br>*S. aureus* : 2 (6.5)<br>*E. faecalis* : 2 (6.5)<br>*K. pneumoniae* : 2 (6.5)<br>*S. pneumoniae* : 1 (3.2)<br>*S. capitis* : 1 (3.2)<br>*E. avium* : 1 (3.2)<br>*P. mirabilis* : 1 (3.2)<br>*S. marcescens* : 1 (3.2)<br>*E. cloacae* : 1 (3.2)<br>*E. aerogenes* : 1 (3.2)<br>*Lactobacillus* spp : 1 (3.2) | *S. aureus* : 6 (26.1)<br>*E. coli* : 5 (21.7)<br>*E. faecalis* : 2 (8.7)<br>*E. cloacae* : 2 (8.7)<br>*P. aerguinosa* : 2 (8.7)<br>*S. epidermidis* : 2 (8.7)<br>*E. avium* : 2 (8.7)<br>*K. pneumoniae* : 1 (4.3)<br>*M. morgagnii* : 1 (4.3)<br>*C. freundii* : 1 (4.3)<br>*B. fragilis* : 1 (4.3) |
| *Klebsiella pneumoniae* | *E. cloacae* : 4 (25.0)<br>*P. aeruginosa* : 3 (18.8)<br>*E. faecalis* : 2(12.5)<br>*S. epidermidis* : 1 (6.3)<br>*S. aureus* : 1 (6.3)<br>*E. coli* : 1 (6.3)<br>*K. pneumoniae* : 1 (6.3)<br>*M. morgagnii* : 1 (6.3)<br>*S. marcescens* : 1 (6.3)<br>*Acinetobacter lwoffii* : 1 (6.3) | *E. cloacae* : 3 (37.5)<br>*S. aureus* : 1 (12.5)<br>*E. faecalis* : 1 (12.5)<br>*E. coli* : 1 (12.5)<br>*M. morgagnii* : 1 (12.5)<br>*P. aeruginosa* : 1 (12.5) | *P. aeruginosa* : 2 (25)<br>S. epidermidis: 1 (12.5)<br>E. faecalis: 1 (12.5)<br>K. pneumoniae: 1 (12.5)<br>S. marcescens: 1 (12.5)<br>E. cloacae: 1 (12.5)<br>A. lwoffii : 1 (12.5) |
| *Klebsiella oxytoca* | *E. cloacae* : 1 (25.0)<br>*K. oxytoca* : 1 (25.0)<br>*E. faecalis* : 1 (25.0)<br>*P. aeruginosa* : 1 (25.0) | - | *E. cloacae* : 1 (25.0)<br>*K. oxytoca* : 1 (25.0)<br>*E. faecalis* : 1 (25.0)<br>*P. aeruginosa* : 1 (25.0) |
| *Enterobacter cloacae* | *P. aeruginosa* :3 (15.8)<br>*Streptococcus spp.* : 3 (15.8)<br>*S. epidermidis*: 2 (10.5)<br>*E. faecalis* : 2 (10.5)<br>*E. coli* : 2 (10.5)<br>*S.marcescens* : 2 (10.5)<br>*E. faecium* : 1 (5.3)<br>*P. mirabilis* : 1 (5.3)<br>*E. cloacae* : 1 (5.3)<br>*H. alvei* : 1 (5.3) | *S. epidermidis* : 1 (11.1)<br>*E. faecalis* : 1 (11.1)<br>*E. faecium* : 1 (11.1)<br>*P. mirabilis* : 1 (11.1)<br>*E. coli* : 1 (11.1)<br>*H. alvei* : 1 (11.1)<br>*P. aeruginosa* : 1 (11.1)<br>*B. fragilis* : 1 (11.1) | *Streptococcus* spp. : 3 (30.0)<br>*P. aeruginosa* : 2 (20.0)<br>*S. marcescens* : 2 (20.0)<br>*E. coli*: 1 (10.0)<br>*S. marcescens :* 1 (10.0)<br>*E. cloacae :* 1 (10.0) |

| | |
|---|---|
| | *Bacteroïdes fragilis* : 1 (5.3) |

Supplementary figure 1:

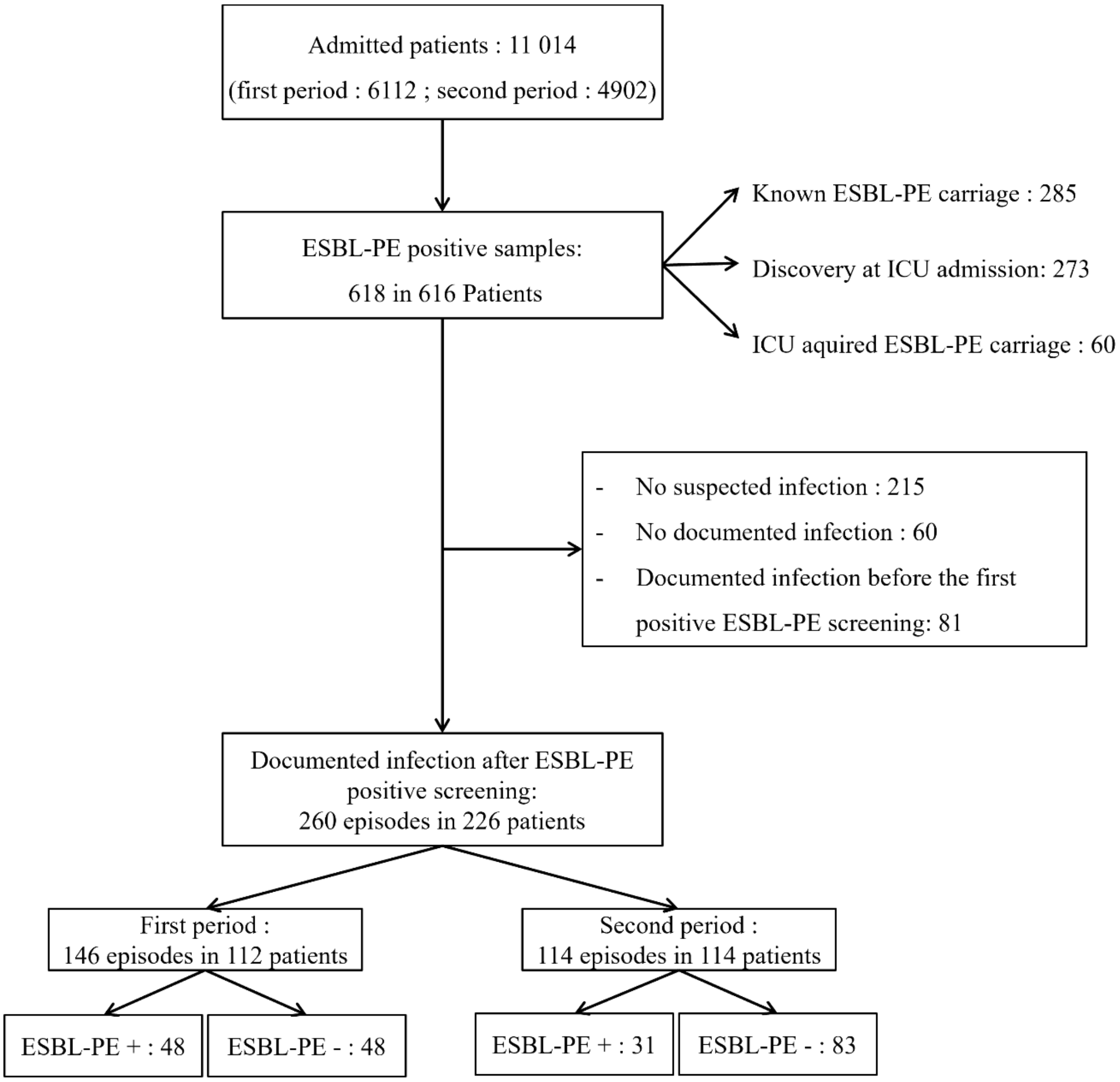

Figure 1 : Flow chart
All patients included during the two study periods are shown in the flow chart. The two periods are distinguished only in terms of the total number of ICU admissions during each period.
ESBL-PE : Extended-spectrum β-lactamases-producing *Enterobacterales;* ICU : Intensive care unit
ESBL-PE + : infection involving and ESBL-EP
ESBL-PE - : infection not involving and ESBL-EP

Supplementary figure 2:

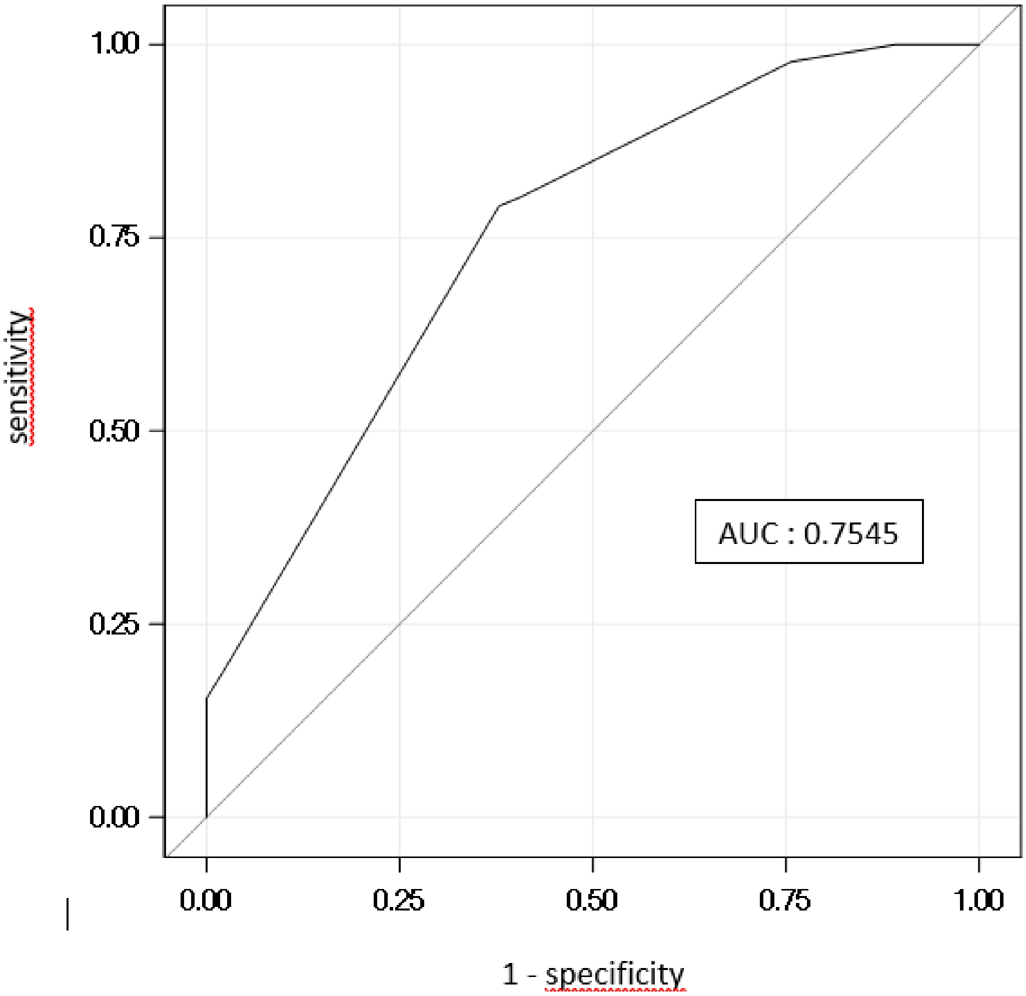
